# Comparative effectiveness of biologics in patients with rheumatoid arthritis stratified by body mass index and sex: a cohort study in SCQM

**DOI:** 10.1101/2022.09.30.22280396

**Authors:** Enriqueta Vallejo-Yagüe, Theresa Burkard, Axel Finckh, Andrea M. Burden, the clinicians and patients of the Swiss Clinical Quality Management Program

## Abstract

**Background:** Obesity is associated with lower treatment response in patients with rheumatoid arthritis (RA). Among obese patients, abatacept was suggested as a preferable option to tumour necrosis factor alpha (TNF) inhibitors. Sex and gender differences in RA were described.

**Objectives:** To assess the comparative effectiveness of etanercept, infliximab, and abatacept, compared to adalimumab, in patients with RA stratified by body mass index (BMI) and sex.

**Methods:** Observational cohort study in the Swiss Clinical Quality Management in Rheumatic Diseases (SCQM) registry (1997-2019). RA patients were classified in BMI-based cohorts: obese, overweight, and normal weight. Each BMI cohort was studied overall and stratified by sex. The study outcome was remission within 12-months, defined as a disease activity score (DAS28) <2.6. Missingness was addressed using confounder-adjusted response rate with attrition correction (CARRAC). Logistic regression compared the effectiveness of etanercept, infliximab, and abatacept versus adalimumab.

**Results:** The study included 443 obese, 829 overweight, and 1243 normal weight RA patients. Across the BMI cohorts, there were no significant differences in the odds of remission at ≤12-months for the study drugs compared to adalimumab. However, among females, an inverse effect for infliximab was found, whereby overweight patients had higher odds of remission, while obese patients had lower odds of remission, compared to the respective adalimumab users.

**Conclusions:** Despite the previous hypothesis, treatment with abatacept showed similar odds of remission compared to adalimumab in all BMI cohorts. Conversely, compared to adalimumab, infliximab performed better in overweight female patients but worse in female patients with obesity. However, further validation is needed.

## INTRODUCTION

Rheumatoid arthritis (RA) is a chronic autoimmune disease, primarily characterised by joint damage, which can lead to disability.^1,2^ Its pathogenesis and clinical presentation may vary between individuals and disease stages.^1^ Following failure to achieve the therapeutic target with conventional synthetic disease-modifying anti-rheumatic drugs (csDMARD), the European Alliance of Associations for Rheumatology (EULAR) recommends adding a biologic or targeted synthetic disease-modifying anti-rheumatic drug (b/tsDMARD).^3^ Supported by a recent systematic review,^4^ the current EULAR guidelines have no preference for specific b/tsDMARD due to similar efficacy.^3^

Despite the advances in the treatment of RA and the availability of several b/tsDMARDs, up to 60% of patients will either not respond or lose response to therapy over time.^5–8^ Thus, evidence-based decision on the optimal b/tsDMARD for each patient remains challenging. This is specifically important for RA patients with high body mass index (BMI) since obesity has been associated with worse disease activity and disease management in patients with RA,^9–14^ and the prevalence of obesity was reported higher among RA cohorts compared to the reference populations.^15,16^ There are hypotheses to explain the reduced therapeutic response in patients with obesity. First, obesity is a low-grade systemic inflammatory condition,^17^ which may share a common pathological pathway with immune-mediated diseases. Second, body weight can affect the drug’s volume of distribution^18^. Third, the probability of developing anti-drug-antibodies (ADAbs) grows when body weight increases.^19^ And fourth, obesity may affect and be affected by socially-constructed norms and behaviours with an impact on clinical management (e.g., weight stigma associated with less exercise^20^).

While previous studies have shown that obesity is associated with a detrimental response to tumour necrosis factor alpha (TNF) inhibitors,^10,14,21^ it has been suggested that high BMI does not influence the response to the non-TNF biologic abatacept.^22–24^ However, these studies assessed the impact of obesity on the treatment response solely among users of abatacept,^22–24^ and often had small sample sizes.^22,24^ Thus, it remains of interest to study the comparative effectiveness of TNF inhibitors versus abatacept in RA patients with obesity. Additionally, although similar effectiveness was suggested across TNF inhibitors in the general RA population,^25^ it is unclear if this is the case in every BMI group.

Sex and gender-based differences in RA were described, including differences in the immune system, drug pharmacokinetics, treatment response rates, and immunity.^26–28^ Therefore, sex-stratified analyses are of interest.

Thus, we decided to perform a comparative effectiveness analysis among RA patients who were new-users of biologics in the Swiss Clinical Quality Management in Rheumatic Diseases (SCQM) cohort, stratified by BMI category and, secondarily, by sex.

## METHODS

### Data source and study design

An observational cohort study in the SCQM registry from 1st January 1997 to 31st July 2019. The SCQM includes routinely collected data from rheumatology visits and patient-reported outcomes, including patient demographics, lifestyle habits, clinical endpoints, anti-rheumatic medication (with start and stop dates), patient-reported outcomes, and health standardised surveys.^16,29^ More details have been described elsewhere.^16^

### Study population

The study included adult (>18 years) RA patients registered in SCQM, who started adalimumab, etanercept, infliximab, or abatacept as their first b/tsDMARD between 1^st^ January 1997 and 31^st^ July 2018. Patients were stratified by BMI category at the start of treatment (index date). BMI categories were obese (BMI≥30 kg/m^2^), overweight (≥25 and <30 kg/m^2^), and normal weight (BMI ≥18.5 and <25 kg/m^2^). Each BMI group was studied as an independent cohort, overall and stratified by sex (i.e., female, male). We excluded patients without a baseline BMI record and underweight patients (BMI <18.5 kg/m^2^).

### Exposure

The study exposure was the patient’s first b/tsDMARD, including etanercept, infliximab, and abatacept, compared to adalimumab.

### Outcome and follow-up

The primary outcome was clinical response during the treatment course with a maximum follow-up of 12-months. Clinical response was primarily defined as 28-joint Disease Activity Score (DAS28) remission (DAS28<2.6), which was calculated using the erythrocyte sedimentation rate (ESR, DAS28-ESR). Secondarily, clinical response was also assessed as DAS28 low disease activity (LDA), defined as DAS28<3.2; and Rheumatoid Arthritis Disease Activity Index-Five (RADAI-5) remission, defined as RADAI-5≤1.4. Treatment course was assessed using drug-specific extended time-windows after treatment stop. These were 42 days for adalimumab, 30 days for etanercept, 90 days for infliximab, and 60 or 30 days for i.v. and s.c. abatacept, respectively. Additionally, a permissible gap of up to 1-month between stop and restart of the same treatment was accepted as treatment continuation. A schematic representation of the follow-up for the primary outcome can be seen in **Supplementary Figure S1**.

Additional secondary outcomes were the median change (Δ, delta) in unidimensional parameters between baseline and the best respective measurement during follow-up as described above. These included ΔESR, delta C-reactive protein (ΔCRP), delta tender joint counts (ΔTJC28), and delta swollen joint counts (ΔSJC28). Here, median values <0 reflect improvement and reduction of the respective values.

Following recent recommendations from EULAR,^30,31^ missing information on primary and secondary outcomes was addressed using the confounder-adjusted response rate with attrition correction (CARRAC).^31^ This consisted of multiple imputation by chain equation (MICE) that included baseline variables, treatment duration, and reason for treatment discontinuation. Additionally, missingness for the clinical response outcomes was also addressed in two other manners as sensitivity analyses: first, assuming that lack of information on outcome during follow-up was equivalent to not-achieving the outcome (MOIAN, Missing Outcome Information Assumed as No); and second, excluding patients who miss this information on outcome during follow-up (EPMOI, Excluding Patients Missing Outcome Information).

The tertiary outcome was treatment survival with a maximum follow-up of 5-years, overall and stratified by the reason for treatment stop adverse event(s), or remission, as recorded by the clinician. For this, we used the record of treatment stop without additional time extension and accepted ≤1-month gaps between stop and re-start of the same biologic as treatment continuation. Treatment stop was defined by a record of stop or by the start of a new b/tsDMARD. Otherwise, patients were censored at the time of stopping their participation at SCQM, at the end of the study period (31^st^ July 2019), or 3-months after a visit with no subsequent visits for >2-years.

### Covariates

Patient baseline characteristics were collected at the index date or within pre-defined lookback windows. Information on patient demographics, disease duration (time from RA diagnosis), seropositivity, swollen and tender joint counts (SJC28, TJC28), physician global disease activity (GDA), and body weight were collected within the 6-months prior index date. Inflammatory markers (ESR, CRP), disease activity score, and the Health Assessment Questionnaire (HAQ) were collected within the 3-months prior index date. Information on smoking (ever smoker), body height, and comorbidities were collected with an ever-before look-back window, except for records on fractures/surgeries/musculoskeletal system, which were collected within the 6-months prior index date. Information on pregnancy or breastfeeding was collected with a 12-month look-back window. Information on rheumatic medication was collected at the index date, including conventional synthetic disease-modifying anti-rheumatic drug (csDMARD) use, steroid use, and type of b/tsDMARD.

### Statistical analysis

The obese, overweight, and normal weight groups were addressed as three distinctive cohorts. Patient baseline characteristics for each study cohort were described stratified by the exposure drug. The etanercept, infliximab, and abatacept groups were compared to the adalimumab group using chi-squared test for categorical variables and t-test for continuous variables. For these tests, missing values did not function as a grouping variable.

CARRAC was performed prior to the analysis of the clinical response outcomes and the change in unidimensional parameters. We performed 60 imputations on an outcome and cohort basis. We visually assessed the convergence of the imputations by mean and variance changes and addressed the overlapping of the distribution of continuous variables with density plots. Information on included variables and methods used in the imputations are described in **Supplementary Table S1**, and an example of visual assessment of the imputation of DAS28-ESR for the primary outcome (DAS28-remission) is depicted in **Supplementary Figure S2**.

Comparative effectiveness of the study drugs for the clinical response outcomes was assessed using logistic regression, with adalimumab as the reference group. Following the CARRAC, logistic regression was performed in each imputed dataset, and the results were subsequently pooled into a single estimate according to Rubin’s rules. This regression was conducted, first, adjusting for age and sex, and second, adjusting for age, sex, index year, baseline DAS28, csDMARD at index, and steroid use at index. Sensitivity analyses were performed by MOIAN and EPMOI, followed by logistic regression calculating age and sex-adjusted odds ratio (OR).

Change in individual parameters (ΔESR, ΔCRP, TJC28, ΔSJC28) was described using the median and interquartile range (IQR) and the Kruskal-Wallis test compared between the exposure drugs, using adalimumab as reference. Lastly, treatment survival was investigated with Kaplan-Meier curves for each cohort overall and stratified by reason of treatment stop (adverse event(s); remission) as recorded by the clinician. Treatment survival across drugs was compared using the log-rank test.

All analyses were independently performed for each BMI cohort (obese, overweight, and normal weight) overall and stratified by sex (female; male). The statistical analyses were performed with the R software, version 3.5.2.^33^

## RESULTS

The study included 2515 RA patients, among whom 443 (17.6%), 829 (33.0%), and 1243 (49.4%) were included in the obese, overweight, and normal weight cohorts, respectively (**Supplementary Figure S3**). The number of users of each study drug and their percentage within the study sub-cohorts (BMI cohorts stratified by sex) is depicted in **Figure 1**. The most commonly prescribed drugs were adalimumab and etanercept, followed by infliximab and abatacept. An increased use of abatacept was observed in the obese versus the normal weight cohort, especially among male patients.

**Figure 1.**
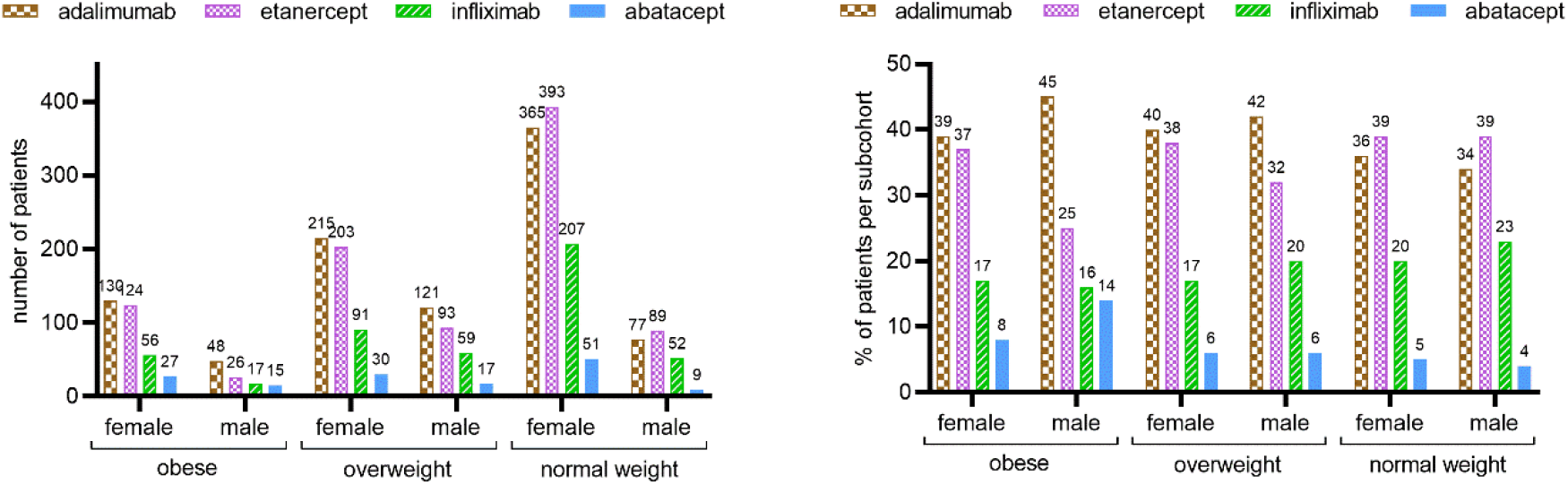
Number (left) and frequency (right) of patients using each study drug within each study sub-cohort (BMI cohorts stratified by sex). The number on the columns indicates the number or percentage of patients, respectively.

Baseline characteristics for the obese and overweight cohorts are described in **Table 1**, and additional information is provided in **Supplementary Tables S3** and **S4**. Baseline characteristics for the normal weight cohort are described in **Supplementary Table S5**. In every BMI cohort, the median year of index date generally differed between the study drugs, with infliximab having the earliest and abatacept the latest. Etanercept users were very similar to adalimumab users in all BMI categories but had a significantly lower percentage of csDMARD use at index date in the obese and normal weight cohorts. Compared to the adalimumab group, infliximab users had significantly more frequent use of prednisone at index in every BMI cohort, significantly more frequent use of csDMARD at index, worse HAQ, and more frequent depression/anxiety in the overweight and normal weight cohorts. In comparison to the adalimumab group, abatacept users were more frequently current or ever smokers in the overweight and obese cohorts and generally had more frequent history of hyperlipidemia and cardiac/cardiovascular event/disease and a tendency for more frequent diabetes. The characteristics stratified by BMI and sex are provided in **Supplementary Tables S6-S11**.

**Table 1.** Obese and overweight cohorts, patient characteristics at baseline, stratified by first b/tsDMARD adalimumab, etanercept, infliximab, and abatacept.

|  | Adalimumab |  | Etanercept |  | Infliximab |  | Abatacept |  |
| --- | --- | --- | --- | --- | --- | --- | --- | --- |
| Obese cohort | (n=178) |  | (n=150) | p-value | (n=73) | p-value | (n=42) | p-value |
| Women (%) | 130 (73.0) |  | 124 (82.7) | 0.052 | 56 (76.7) | 0.656 | 27 (64.3) | 0.348 |
| Age index (mean (SD)) | 56.60 (11.99) |  | 56.98 (11.63) | 0.777 | 55.86 (10.77) | 0.644 | 59.27 (10.00) | 0.183 |
| RA duration, years (mean (SD)) | 6.72 (8.67) |  | 7.75 (8.45) | 0.285 | 7.92 (8.61) | 0.331 | 5.80 (7.42) | 0.531 |
| Year of index date (mean (SD)) | 2007.85 (3.79) |  | 2007.53 (4.56) | 0.487 | 2005.68 (4.02) | <0.001 | 2013.19 (2.52) | <0.001 |
| BMI kg/m<sup>2</sup> (mean (SD)) | 33.90 (3.86) |  | 33.79 (3.85) | 0.794 | 33.65 (3.09) | 0.621 | 33.91 (3.99) | 0.987 |
| Ever smoker (%) | 49 (27.5) |  | 41 (27.3) | 1.000 | 11 (15.1) | 0.053 | 22 (52.4) | 0.004 |
| csDMARD at index date (%) | 134 (75.3) |  | 96 (64.0) | 0.036 | 60 (82.2) | 0.307 | 33 (78.6) | 0.804 |
| Prednisone at index date (%) | 66 (37.1) |  | 60 (40.0) | 0.669 | 41 (56.2) | 0.008 | 18 (42.9) | 0.605 |
| Seropositive <sup>a</sup> (%) | 129 (72.5) |  | 102 (68.0) | 0.691 | 56 (76.7) | 0.537 | 32 (76.2) | 0.765 |
| ESR (mean (SD)) | 24.22 (18.46) |  | 23.73 (16.09) | 0.809 | 29.43 (18.87) | 0.053 | 22.00 (18.05) | 0.518 |
| CRP (mean (SD)) | 1.44 (1.20) |  | 1.37 (1.42) | 0.765 | 1.38 (1.08) | 0.842 | 1.06 (0.97) | 0.106 |
| Tender joint counts 28 (mean (SD)) | 7.25 (6.92) |  | 7.86 (6.75) | 0.432 | 8.14 (7.20) | 0.367 | 7.19 (6.66) | 0.963 |
| Swollen joint counts 28 (mean (SD)) | 6.75 (5.72) |  | 6.63 (5.79) | 0.865 | 7.38 (6.37) | 0.449 | 5.73 (4.85) | 0.317 |
| Physician GDA (mean (SD)) | 4.94 (1.85) |  | 4.93 (1.95) | 0.950 | 5.29 (2.13) | 0.344 | 4.09 (1.99) | 0.024 |
| DAS28-ESR (mean (SD)) | 4.40 (1.42) |  | 4.54 (1.32) | 0.406 | 4.72 (1.40) | 0.118 | 4.18 (1.46) | 0.399 |
| RADAI-5 (mean (SD)) | 4.91 (2.04) |  | 5.33 (2.18) | 0.107 | 5.15 (2.12) | 0.425 | 4.35 (2.36) | 0.209 |
| HAQ (mean (SD)) | 1.21 (0.74) |  | 1.32 (0.73) | 0.235 | 1.38 (0.77) | 0.115 | 0.95 (0.70) | 0.093 |
| Osteoporosis <sup>b</sup> | 24 (13.5) |  | 23 (15.3) | 0.750 | 11 (15.1) | 0.898 | 4 (9.5) | 0.663 |
| Other rheumatological disease(s) <sup>c</sup> | 61 (34.3) |  | 62 (41.3) | 0.229 | 25 (34.2) | 1.000 | 8 (19.0) | 0.084 |
| Psoriasis | 2 (1.1) |  | 2 (1.3) | 1.000 | 1 (1.4) | 1.000 | 0 (0.0) | 1.000 |
| Hyperlipidemia | 15 (8.4) |  | 11 (7.3) | 0.873 | 5 (6.8) | 0.871 | 10 (23.8) | 0.011 |
| Cardiac/cardiovascular event/disease <sup>d</sup> | 84 (47.2) |  | 84 (56.0) | 0.139 | 32 (43.8) | 0.730 | 30 (71.4) | 0.008 |
| Depression/anxiety <sup>e</sup> | 25 (14.0) |  | 31 (20.7) | 0.150 | 12 (16.4) | 0.772 | 7 (16.7) | 0.849 |
| Diabetes | 17 (9.6) |  | 23 (15.3) | 0.154 | 9 (12.3) | 0.669 | 8 (19.0) | 0.140 |
| Fractures, surgeries, musc. system | 15 (8.4) |  | 8 (5.3) | 0.381 | 7 (9.6) | 0.960 | 3 (7.1) | 1.000 |
| **Overweight cohort** | **(n=336)** |  | **(n=296)** | **p-value** | **(n=150)** | **p-value** | **(n=47)** | **p-value** |
| Women (%) | 215 (64.0) |  | 203 (68.6) | 0.257 | 91 (60.7) | 0.549 | 30 (63.8) | 1.000 |
| Age index (mean (SD)) | 57.28 (12.52) |  | 57.81 (12.32) | 0.589 | 56.87 (11.35) | 0.734 | 62.99 (10.81) | 0.003 |
| RA duration, years (mean (SD)) | 7.10 (7.76) |  | 8.45 (9.21) | 0.050 | 7.66 (8.08) | 0.477 | 6.60 (8.45) | 0.690 |
| Year of index date (mean (SD)) | 2007.68 (3.56) |  | 2006.72 (5.02) | 0.005 | 2005.93 (4.69) | <0.001 | 2012.32 (2.60) | <0.001 |
| BMI kg/m<sup>2</sup> (mean (SD)) | 27.16 (1.35) |  | 27.21 (1.33) | 0.643 | 27.13 (1.37) | 0.829 | 27.19 (1.41) | 0.885 |
| Ever smoker (%) | 94 (28.0) |  | 76 (25.7) | 0.575 | 43 (28.7) | 0.962 | 22 (46.8) | 0.014 |
| csDMARD at index date (%) | 226 (67.3) |  | 189 (63.9) | 0.414 | 126 (84.0) | <0.001 | 34 (72.3) | 0.595 |
| Prednisone at index date (%) | 134 (39.9) |  | 133 (44.9) | 0.229 | 84 (56.0) | 0.001 | 13 (27.7) | 0.146 |
| Seropositive <sup>a</sup> (%) | 244 (72.6) |  | 211 (71.3) | 0.679 | 119 (79.3) | 0.731 | 37 (78.7) | 0.697 |
| ESR (mean (SD)) | 24.82 (19.57) |  | 25.87 (22.59) | 0.555 | 25.44 (21.41) | 0.766 | 28.92 (18.07) | 0.221 |
| CRP (mean (SD)) | 2.12 (3.25) |  | 1.71 (3.10) | 0.315 | 1.18 (1.68) | 0.057 | 1.30 (1.48) | 0.121 |
| Tender joint counts 28 (mean (SD)) | 7.55 (6.90) |  | 7.95 (7.69) | 0.507 | 7.14 (6.85) | 0.552 | 6.00 (5.88) | 0.161 |
| Swollen joint counts 28 (mean (SD)) | 6.90 (5.51) |  | 6.68 (5.93) | 0.639 | 7.92 (5.98) | 0.077 | 5.82 (5.18) | 0.219 |
| Physician GDA (mean (SD)) | 4.92 (2.27) |  | 4.90 (2.14) | 0.907 | 5.56 (2.04) | 0.033 | 4.28 (1.89) | 0.091 |
| DAS28-ESR (mean (SD)) | 4.47 (1.34) |  | 4.42 (1.50) | 0.690 | 4.42 (1.48) | 0.747 | 4.49 (1.15) | 0.933 |
| RADAI-5 (mean (SD)) | 4.84 (2.08) |  | 5.01 (2.17) | 0.358 | 4.98 (2.18) | 0.528 | 4.87 (2.35) | 0.935 |
| HAQ (mean (SD)) | 1.08 (0.70) |  | 1.13 (0.74) | 0.408 | 1.24 (0.72) | 0.030 | 0.94 (0.69) | 0.272 |
| Osteoporosis <sup>b</sup> | 59 (17.6) |  | 61 (20.6) | 0.382 | 33 (22.0) | 0.303 | 11 (23.4) | 0.442 |
| Other rheumatological disease(s) <sup>c</sup> | 101 (30.1) |  | 97 (32.8) | 0.517 | 45 (30.0) | 1.000 | 13 (27.7) | 0.868 |
| Psoriasis | 2 (0.6) |  | 3 (1.0) | 0.887 | 0 (0.0) | 0.857 | 1 (2.1) | 0.816 |
| Hyperlipidemia | 16 (4.8) |  | 25 (8.4) | 0.086 | 8 (5.3) | 0.967 | 7 (14.9) | 0.016 |
| Cardiac/cardiovascular event/disease <sup>d</sup> | 123 (36.6) |  | 117 (39.5) | 0.501 | 58 (38.7) | 0.740 | 21 (44.7) | 0.363 |
| Depression/anxiety <sup>e</sup> | 31 (9.2) |  | 42 (14.2) | 0.068 | 27 (18.0) | 0.009 | 2 (4.3) | 0.390 |
| Diabetes | 26 (7.7) |  | 19 (6.4) | 0.625 | 8 (5.3) | 0.443 | 8 (17.0) | 0.068 |
| Fractures, surgeries, musc. system | 26 (7.7) |  | 34 (11.5) | 0.142 | 13 (8.7) | 0.867 | 2 (4.3) | 0.575 |

Values are the number and column percentage, unless otherwise specified. Significance tests compare each drug of interest to adalimumab, using chi-squared test for categorical variables, and t-test for continuous variables. For these tests, missing values did not function as a grouping variable.
Abbreviations: RA rheumatoid arthritis; BMI body mass index; csDMARD conventional synthetic disease modifying anti-rheumatic drug; ESR erythrocyte sedimentation rate; CRP C-reactive protein; GDA global disease activity; DAS28-ESR 28-joint Disease Activity Score using erythrocyte sedimentation rate; RADAI-5 Rheumatoid Arthritis Disease Activity Index-Five; HAQ Health Assessment Questionnaire; musc. musculoskeletal.
<sup>a</sup> Seropositivity was calculated using both rheumatoid factor (RF) and anti-cyclic citrullinated peptide (anti-CCP) antibodies; <sup>b</sup> Osteoporosis includes osteoporosis record or medication with bisphosphonates, denosumab, or teriparatide; <sup>c</sup> Other rheumatological disease includes gout, lupus, osteoarthritis, Sjogren syndrome, degenerative spine disease, degenerative spondylopathy, other connective tissue disease, other rheumatological disease; <sup>d</sup> Cardiac/cardiovascular event/disease includes myocardial infarction, heart infarct, heart failure, heart insufficiency, cardiac insufficiency, coronary heart disease, coronary cardiac disease, heart problem, heart disease, angina pectoris, rhythm disorder, artery intervention, stroke transient ischemic attack, cerebrovascular disease, deep venous thrombosis, peripheral vascular disease, pulmonary embolism, blood thinners, hypertension, hypotension, other cardiovascular disease, and medication with platelet aggregation inhibitors, antihypertensives, or statins; <sup>e</sup> Depression/anxiety includes record of the disease or medication with antidepressants.

**Table 2** provides the results from the comparative effectiveness analysis for the clinical response outcomes (DAS28-remission; DAS28-LDA; RADAI-5-remission) in the overall BMI cohorts using CARRAC and MOIAN. The respective EPMOI analyses are presented in **Supplementary Table S12**. In the overall BMI cohorts, no significant differences were identified across the study drugs compared to adalimumab, with only one exception. In overweight patients, etanercept was associated with a reduced odds of achieving RADAI-5 remission (ORadj 0.44, 95%CI 0.22-0.90). This finding was consistent between both the sex- and age-adjusted model and the full-adjusted model, as well as consistent across the CARRAC, MOIAN, and EPMOI analyses.

**Table 2.**
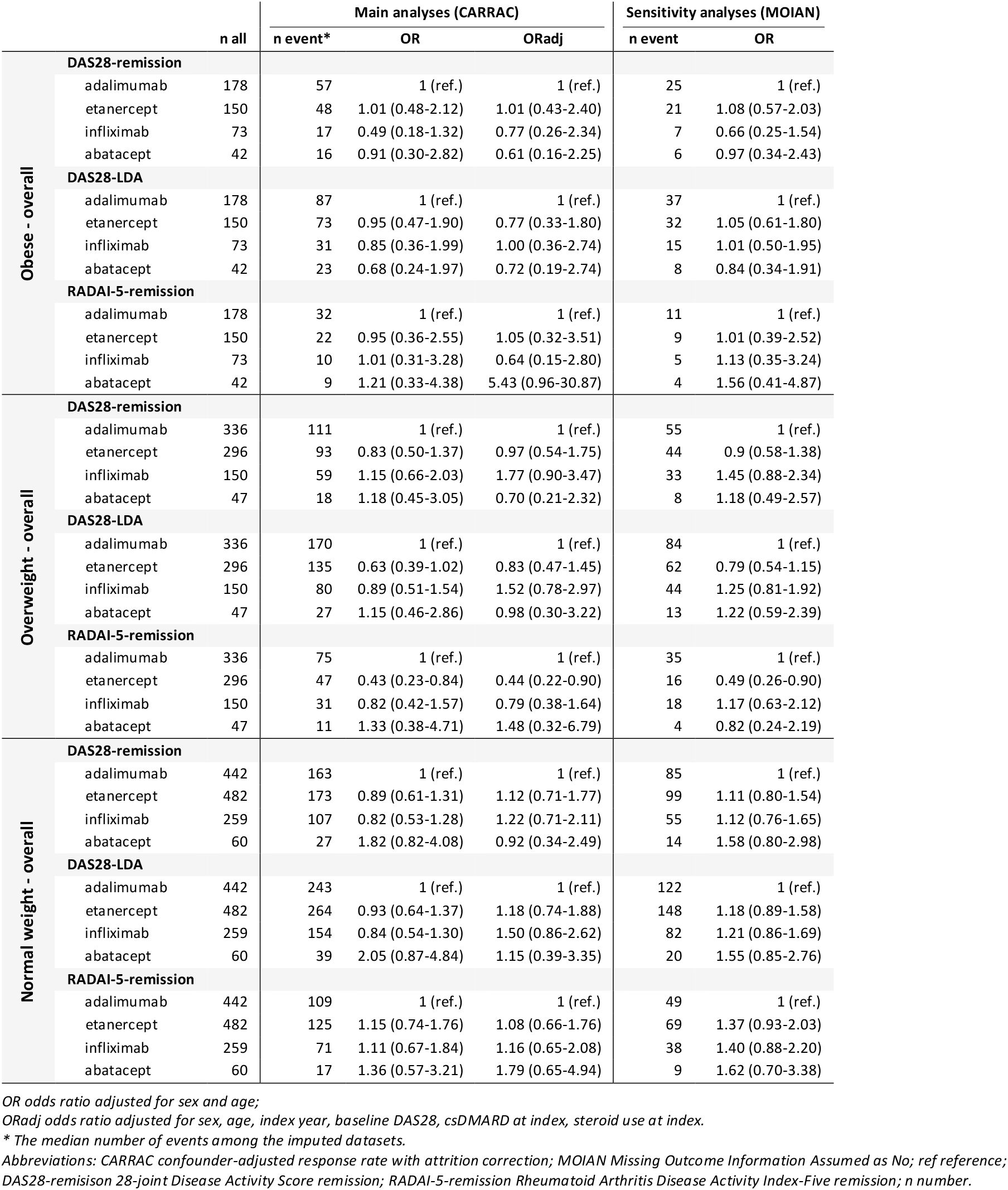
Comparative effectiveness analyses.

**Table 3** presents the analysis among females, including CARRAC and MOIAN. Additionally, EPMOI analyses are presented in **Supplementary Table S13**. Obese female patients treated with infliximab had lower odds of achieving DAS28-remission in the CARRAC age-adjusted model (OR 0.20, 95%CI 0.04-0.96), MOIAN (OR 0.26, 95%CI 0.04-0.97), and EPMOI (OR 0.20, 95%CI 0.03-0.79) analyses. However, this effect was not significant in the CARRAC full-adjusted analysis (ORadj 0.27, 95%CI 0.05-1.41). Conversely, in the overweight female cohort, higher odds of remission were observed with infliximab (ORadj 2.47, 95%CI 1.06-5.78). This effect was observed in the CARRAC full-adjusted and MOIAN analyses but not in the EPMOI analyses.

**Table 3.**
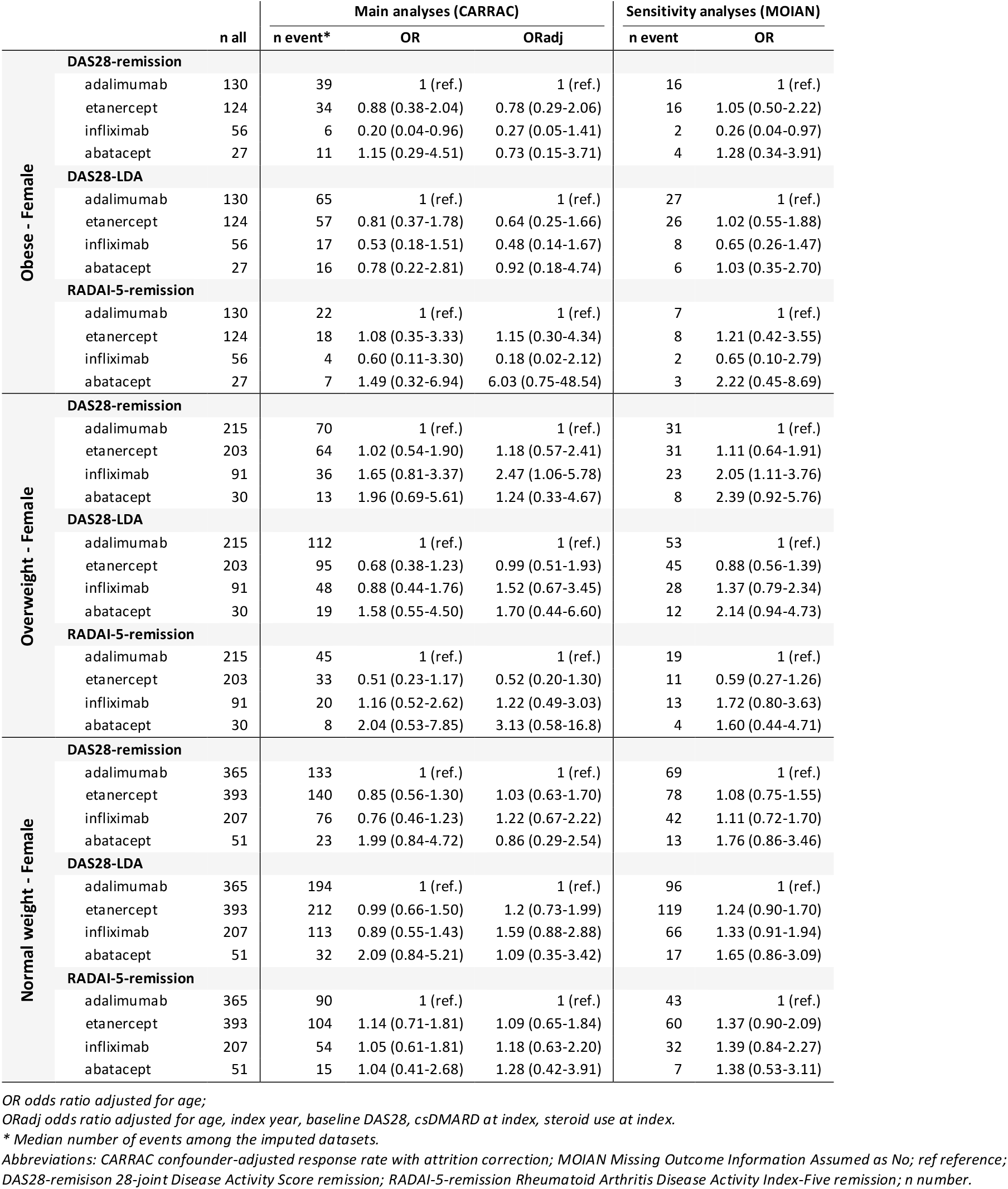
Comparative effectiveness analyses. Female cohorts.

The stratification among males is provided in **Table 4**, and the EPMOI analyses are provided in **Supplementary Table S13**. Similar to the overall analysis, the overweight male users of etanercept had reduced odds of achieving RADAI-5 compared to adalimumab users. This was observed in the MOIAN and EPMOI analyses; however, not according to the CARRAC analysis.

**Table 4.**
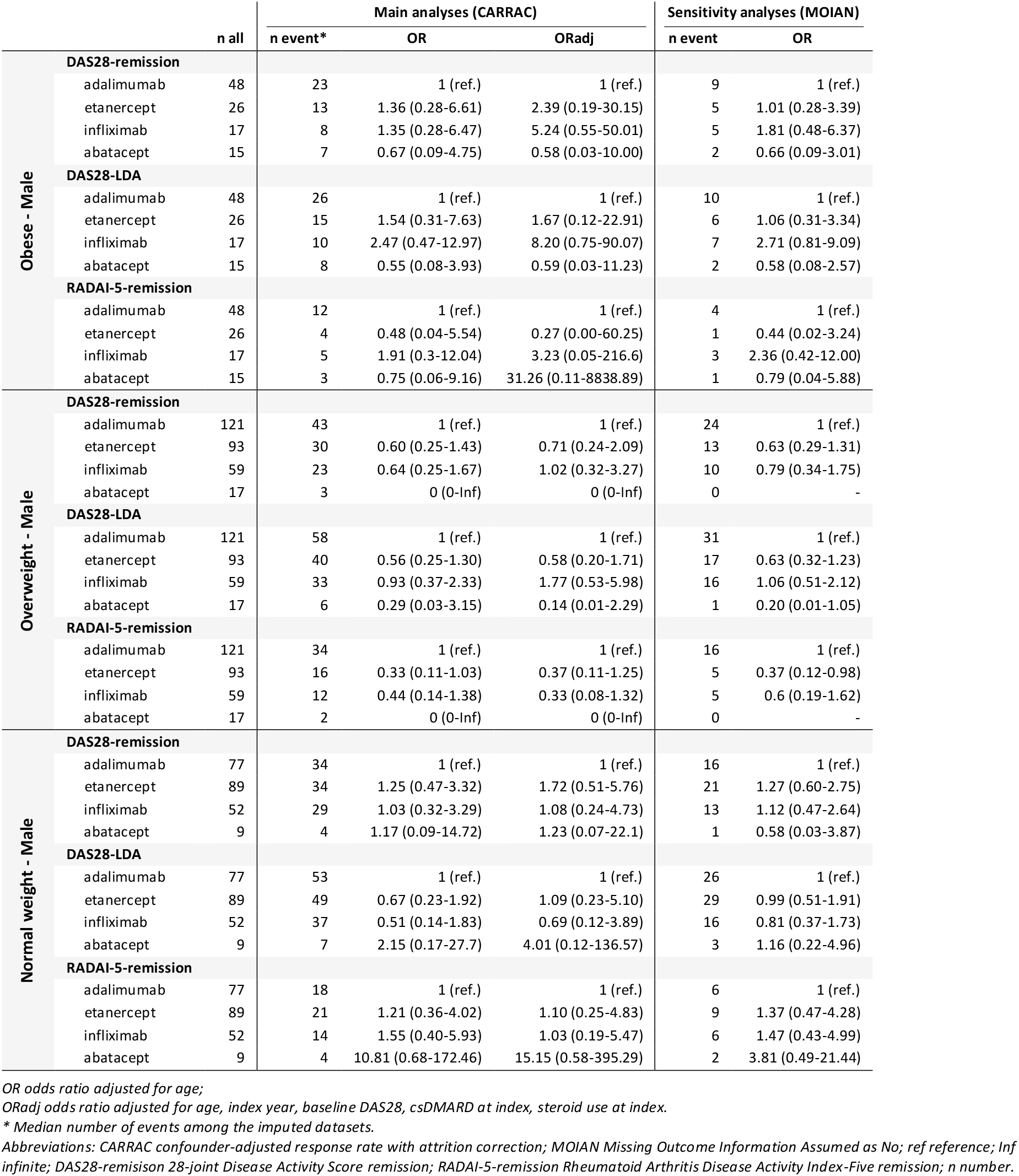
Comparative effectiveness analyses. Male cohorts.

The change in individual parameters is presented in **Table 5**. Among etanercept users, the overweight cohort had a significantly lower reduction (worse improvement) of CRP compared to the respective adalimumab group. This, however, was not significant when stratified by sex. For infliximab users, obese patients had significantly worse improvement on ESR and CRP, yet, in the normal weight cohort, there was a significantly higher improvement in ESR and a tendency for improvement in CRP when compared to adalimumab. The sex-stratified analysis showed that female patients with obesity had significantly worse improvement on ESR, while male obese patients had significantly worse improvement on CRP in comparison to the adalimumab users. Finally, no differences were found between abatacept and adalimumab.

**Table 5.** Median change (delta, Δ) on individual clinical endpoints between baseline and the end of follow-up.

|  |  | Adalimumab | Etanercept | p-value |  | Infliximab | p-value |  | Abatacept | p-value |  |
| --- | --- | --- | --- | --- | --- | --- | --- | --- | --- | --- | --- |
| Overall | **Obese** |  |  |  |  |  |  |  |  |  |  |
| | ΔESR | -3.00 [-11.50, 1.00] | -4.50 [-16.00, 1.00] | 0.596 | | -0.50 [-5.00, 14.75] | 0.044 | | -1.00 [-8.00, 2.00] | 0.574 | |
| | ΔCRP | -0.20 [-1.00, 0.00] | -0.45 [-0.94, -0.10] | 0.389 | | 0.35 [-0.24, 0.50] | 0.047 | | -0.23 [-0.59, 0.00] | 0.366 | |
| | ΔTJC28 | -3.00 [-6.00, 0.00] | -3.00 [-6.50, -0.50] | 0.668 | | -1.00 [-6.00, 0.00] | 0.642 | | -6.00 [-10.00, -0.75] | 0.131 | |
| | ΔSJC28 | -2.00 [-7.00, -1.00] | -4.00 [-7.00, -0.50] | 0.479 | | -2.00 [-5.50, -0.50] | 0.648 | | -4.00 [-7.00, -2.00] | 0.562 | |
|  | **Overweight** |  |  |  |  |  |  |  |  |  |  |
| | ΔESR | -4.00 [-12.00, 2.00] | -3.00 [-12.00, 2.00] | 0.738 | | -3.50 [-15.25, 2.00] | 0.989 | | -8.00 [-13.00, -5.00] | 0.301 | |
| | ΔCRP | -0.40 [-1.40, 0.00] | 0.00 [-0.50, 0.00] | 0.019 | | -0.28 [-0.73, -0.00] | 0.452 | | -0.35 [-0.67, 0.00] | 0.435 | |
| | ΔTJC28 | -3.00 [-6.75, 0.00] | -3.00 [-8.00, 0.00] | 0.713 | | -3.00 [-7.50, 0.00] | 0.882 | | -3.50 [-8.75, -1.00] | 0.627 | |
| | ΔSJC28 | -3.00 [-7.00, -1.00] | -3.00 [-7.00, 0.00] | 0.613 | | -4.00 [-8.50, -1.00] | 0.306 | | -3.00 [-5.50, -0.50] | 0.631 | |
|  | **Normal weight** |  |  |  |  |  |  |  |  |  |  |
| | ΔESR | -3.00 [-14.00, 1.00] | -4.00 [-13.00, 1.00] | 0.942 | | -8.00 [-18.00, 0.00] | 0.040 | | -8.00 [-17.00, 0.00] | 0.290 | |
| | ΔCRP | -0.15 [-0.60, 0.00] | -0.10 [-0.90, 0.00] | 0.808 | | -0.41 [-1.45, 0.00] | 0.173 | | -0.10 [-0.64, 0.00] | 0.701 | |
| | ΔTJC28 | -3.00 [-7.00, 0.00] | -2.00 [-6.00, 0.00] | 0.208 | | -3.00 [-8.50, 0.00] | 0.217 | | -4.00 [-8.00, -2.00] | 0.158 | |
| | ΔSJC28 | -3.00 [-7.00, -0.50] | -3.00 [-6.00, 0.00] | 0.302 | | -5.00 [-9.00, -1.00] | 0.005 | | -3.00 [-7.75, -1.00] | 0.688 | |
| Female | **Obese female** |  |  |  |  |  |  |  |  |  |  |
| | ΔESR | -3.00 [-10.00, 1.00] | -4.00 [-15.00, 1.00] | 0.631 | | 0.50 [-5.00, 19.75] | 0.020 | | 0.00 [-3.00, 3.00] | 0.150 | |
| | ΔCRP | -0.10 [-0.67, 0.00] | -0.40 [-0.80, -0.10] | 0.225 | | 0.05 [-0.79, 0.50] | 0.564 | | -0.23 [-0.40, 0.00] | 0.616 | |
| | ΔTJC28 | -3.00 [-6.00, 0.00] | -3.00 [-7.00, -1.00] | 0.797 | | 0.00 [-4.00, 2.00] | 0.123 | | -6.00 [-10.00, 0.00] | 0.502 | |
| | ΔSJC28 | -2.00 [-7.00, -1.00] | -3.00 [-7.00, 0.00] | 0.851 | | -1.00 [-4.00, 0.00] | 0.123 | | -2.00 [-4.00, 0.00] | 0.738 | |
|  | **Overweight female** |  |  |  |  |  |  |  |  |  |  |
| | ΔESR | -5.00 [-15.00, 1.00] | -2.00 [-10.75, 2.00] | 0.361 | | -4.00 [-15.25, 2.00] | 0.811 | | -8.00 [-12.75, -1.25] | 0.708 | |
| | ΔCRP | -0.30 [-0.90, 0.00] | 0.00 [-0.38, 0.01] | 0.072 | | -0.30 [-0.50, 0.00] | 0.756 | | -0.30 [-0.60, 0.00] | 0.668 | |
| | ΔTJC28 | -2.50 [-7.00, 0.00] | -3.00 [-9.00, 0.00] | 0.649 | | -3.00 [-8.00, 0.00] | 0.794 | | -5.50 [-8.75, -1.00] | 0.508 | |
| | ΔSJC28 | -3.00 [-7.00, -1.00] | -3.00 [-7.00, 0.00] | 0.598 | | -4.00 [-8.00, -1.00] | 0.787 | | -3.00 [-5.75, -1.00] | 0.614 | |
|  | **Normal weight female** |  |  |  |  |  |  |  |  |  |  |
| | ΔESR | -4.00 [-13.75, 1.00] | -3.00 [-12.00, 1.00] | 0.762 | | -10.00 [-18.00, -1.00] | 0.019 | | -8.00 [-17.00, -3.00] | 0.266 | |
| | ΔCRP | -0.10 [-0.50, 0.00] | -0.10 [-0.90, 0.00] | 0.914 | | -0.70 [-1.60, 0.00] | 0.063 | | 0.00 [-0.45, 0.00] | 0.544 | |
| | ΔTJC28 | -3.00 [-7.00, 0.00] | -2.00 [-6.75, 0.00] | 0.470 | | -4.00 [-8.50, 0.00] | 0.271 | | -4.00 [-8.00, -2.00] | 0.283 | |
| | ΔSJC28 | -3.00 [-7.00, 0.00] | -3.00 [-6.00, 0.00] | 0.551 | | -6.00 [-9.00, -1.50] | 0.001 | | -3.00 [-7.75, -1.00] | 0.718 | |
| Male | **Obese male** |  |  |  |  |  |  |  |  |  |  |
| | ΔESR | -2.00 [-17.75, 0.25] | -6.00 [-25.00, 0.00] | 0.509 | | -1.50 [-7.00, -0.25] | 0.774 | | -8.00 [-28.50, -0.50] | 0.422 | |
| | ΔCRP | -1.10 [-1.75, -0.05] | -1.80 [-3.17, -1.05] | 0.357 | | 0.65 [0.43, 0.88] | 0.037 | | -0.35 [-0.69, 0.00] | 0.361 | |
| | ΔTJC28 | -2.50 [-6.75, 0.75] | -2.00 [-4.00, 0.00] | 0.935 | | -4.00 [-8.50, -1.25] | 0.279 | | -6.00 [-12.50, -3.50] | 0.119 | |
| | ΔSJC28 | -2.00 [-6.50, -0.25] | -4.50 [-6.75, -2.25] | 0.221 | | -6.50 [-7.75, -2.00] | 0.253 | | -6.00 [-7.50, -2.00] | 0.203 | |
|  | **Overweight male** |  |  |  |  |  |  |  |  |  |  |
| | ΔESR | -3.50 [-10.25, 2.00] | -5.00 [-16.00, 0.00] | 0.435 | | -3.00 [-12.50, 2.00] | 0.859 | | -8.00 [-16.00, -6.50] | 0.245 | |
| | ΔCRP | -0.61 [-1.63, -0.18] | -0.20 [-0.65, 0.00] | 0.253 | | -0.20 [-0.80, -0.01] | 0.563 | | -0.90 [-2.10, 0.40] | 0.734 | |
| | ΔTJC28 | -3.00 [-4.25, -0.75] | -3.00 [-6.00, 0.00] | 0.951 | | -2.00 [-7.00, 0.00] | 0.913 | | -1.00 [-5.25, -0.25] | 0.543 | |
| | ΔSJC28 | -2.50 [-5.25, -1.00] | -2.00 [-6.00, -0.25] | 0.761 | | -5.00 [-9.00, -1.00] | 0.190 | | -3.00 [-4.00, 0.00] | 0.777 | |
|  | **Normal weight male** |  |  |  |  |  |  |  |  |  |  |
| | ΔESR | -2.50 [-19.00, 0.25] | -8.00 [-14.50, 0.50] | 0.645 | | -2.50 [-10.25, 0.00] | 0.947 | | -12.50 [-30.75, 4.25] | 0.880 | |
| | ΔCRP | -0.20 [-0.85, 0.00] | -0.40 [-0.90, 0.00] | 0.824 | | -0.10 [-0.26, 0.00] | 0.681 | | -0.70 [-1.12, -0.17] | 0.560 | |
| | ΔTJC28 | -3.00 [-6.75, 0.00] | 0.00 [-3.50, 0.50] | 0.169 | | -2.00 [-6.75, 0.00] | 0.583 | | -5.50 [-7.25, -3.50] | 0.282 | |
| | ΔSJC28 | -2.50 [-6.50, -1.00] | -2.00 [-4.00, 0.00] | 0.268 | | -1.00 [-7.00, 0.00] | 0.417 | | -4.50 [-5.75, -2.75] | 0.774 | |

Significance tests compare each drug of interest to adalimumab, using Kruskal-Wallis test.
Abbreviations: n number, sample size; ESR erythrocyte sedimentation rate; CRP C-reactive protein; TJC28 tender joint counts counting 28; SJC28 swollen joint counts counting 28.

Kaplan-Meier curves are depicted in **Supplementary Figure S4**. No differences in drug survival were identified across the study drugs.

## DISCUSSION

This observational cohort study in the SCQM registry included 443 obese, 829 overweight, and 1243 normal weight RA patients treated with adalimumab, etanercept, infliximab, or abatacept as their first b/tsDMARD. In the overall BMI cohorts, similar achievement of DAS28-remission was observed between the studied biologics compared to adalimumab. Results were consistent across various methods and outcomes. However, when stratified by sex, infliximab appeared to perform better among overweight females but worse in obese females in comparison to adalimumab. Additionally, lower odds of achievement of RADAI-5-remission were observed in overweight users of etanercept compared to adalimumab.

Our findings in the overall BMI cohorts were in agreement with published studies on the RA general population.^4,34–36^ For example, a recent observational cohort study of RA patients who were new-users of b/tsDMARDs showed no statistical differences in effectiveness between TNF inhibitors and non-TNF biologics.^34^ Conversely, a study on new-users of adalimumab, etanercept, infliximab, and abatacept reported comparable rates of effectiveness across the study drugs (24%, 28%, 23%, 26%, respectively) but also indicated a lower relative risk of effectiveness for infliximab compared to the other drugs.^37^ In our study, we observed no differences between the clinical response to infliximab and adalimumab in the overall BMI cohorts. However, when stratifying by sex, a contradictory effect was observed in the female cohorts. Infliximab female users with overweight had increased odds of achieving DAS28-remission in comparison to the respective adalimumab group, contrary to the results in the obese female patients, for whom infliximab performed worse than adalimumab. This finding in the obese female users of infliximab was observed in every model but for the CARRAC fully adjusted analyses, in which overfitting is expected due to the very low number of events for this particular finding.

Despite the influence that the body weight has on the volume of distribution of infliximab,^18^ the weight-adjusted dose of this treatment may explain the higher benefit of infliximab versus adalimumab in overweight female patients. However, while one would expect that increasing body fat would have a consistent response, studies among cohorts of RA patients treated with infliximab reported an association between obesity and worse response,^38,39^ consistent with findings from other TNF inhibitors.^10^ Additionally, there may be other factors that influence the low response of infliximab in obese patients. For example, obesity was described as a predictor of hypoalbuminemia,^40^ and serum albumin levels have been inversely associated with the clearance of infliximab.^18^ Thus, lower levels of albumin in obese patients may result in higher clearance of infliximab and, therefore, reduced effectiveness. Additionally, infliximab clearance is not linearly correlated to weight.^19^ Thus, appropriate dose adjustment in overweight patients but altered pharmacokinetics in the presence of highly elevated BMI may explain the conflicting effect observed between the overweight and obese female cohorts. Moreover, our findings on the change of individual parameters suggest that the lower achievement of DAS28-remission in obese female patients with infliximab vs adalimumab may be driven by a significantly lower improvement in inflammation, despite similar improvement in tender and swollen joint counts. This may explain the inconsistency between DAS28-remission and RADAI-5-remission in these patients. RADAI-5 is a patient-driven score, which correlates with tender joint counts, but has a low correlation with ESR.^41^ Thus, despite the validity of RADAI-5 as measurement of disease activity, both scores provide a different assessment of the disease, and the inconsistency between them should not undervalue either. Finally, due to the inflammatory character of obesity, which can result in elevated levels of TNF,^17^ it may be that infliximab does not sufficiently reduce the excess inflammation.

Conversely to the above-discussed results in the female cohort, male patients treated with infliximab and adalimumab had similar odds of achieving remission, irrespectively of their BMI category. This sex difference may be explained by the smaller sample size of the male cohorts. Additionally, sexual dimorphism in body fat may as well play a role. In brief, there are differences in body fat distribution (e.g., males tend to have more visceral adipose tissue, while females have more subcutaneous adipose tissue), adipocyte function, hormonal levels and genetics (with consequent differences in the immune system) between males and females.^42–44^ Thus, this may explain the observed sex differences in response to RA treatment. While further elucidation of this effect in the context of infliximab response is of interest, we consider it beyond the scope of this paper.

Abatacept has been suggested as a preferable drug candidate to treat patients with elevated BMI due to an alternative mode of action. This is supported by the systematic review from Shan and Zhang, which reported reduced odds of response in RA patients with obesity treated with TNF inhibitors but not in patients treated with abatacept.^14^ Four studies have assessed the impact of BMI on the treatment response in RA patients treated with abatacept, all suggesting that BMI does not impact the clinical response to abatacept in RA.^21–24^ In addition to this, the pharmacokinetics of abatacept were consistently described regardless of BMI,^21^ despite abatacept being a lipophilic drug.^22^ This may suggest that the lower response reported in obese patients treated with TNF inhibitors may relate to the mechanistic pathway of these treatments and not solely to their body distribution. For example, body weight was described as a predictor of the formation of ADAbs in RA patients treated with infliximab, potentially explained by the higher TNF-infliximab complexes due to the additional TNF consequence of the adipose tissue.^19^ Therefore, non-TNF biologics open up as potential optimal treatments in obese RA patients. However, while this seemed promising, we did not observe any direct benefit of being treated with abatacept versus adalimumab in any of the study cohorts. This is in agreement with the observed comparable efficacy between abatacept and adalimumab in a head-to-head randomised trial.^45^ Therefore, we trust that current evidence does not justify a superiority of abatacept versus adalimumab in RA patients with obesity.

Regarding etanercept versus adalimumab, the study results showed >50% reduced odds of achieving RADAI-5-remission among etanercept users with overweight in comparison to the respective adalimumab group. However, this effect was not observed for the DAS28 outcomes, and a rationale to explain it is lacking. While this could have been a chance finding, the consistency of this result across the different analysis types (CARRAC, MOIAN, EPMOI) suggests that further investigation is of interest.

### Strengths and limitations

The number of head-to-head trials is increasing,^35^ and studies on the comparative effectiveness of b/tsDMARDs in real-world-setting are limited but rising. However, to our knowledge, this is the first real-world comparative effectiveness observational cohort study on biologics in RA patients stratified by BMI category. Additionally, this is one of the first studies after the very recent recommendation from EULAR to use CARRAC to address missingness during follow-up.^30,31^ Thus, we contribute to the validation of this recommendation while still providing traditional approaches alongside the CARRAC findings.

A limitation of this study is the restriction to only four biologics. This decision was driven by the limited sample size for other b/tsDMARDs due to different times of approval in Switzerland and, importantly, due to former guidelines suggesting TNF inhibitors as preferable first b/tsDMARD choice until 2013.^46,47^ While a prevalence-user design would have enabled to investigate more treatments, we discarded this option to avoid confounding by indication, for example, driven by the expected different response to second-line treatments based on the type of response to the first b/tsDMARD (i.e., primary versus secondary non-response^48^). Finally, although underweight patients were a population of interest, sample size-wise was not feasible to address this research question in these patients.

## CONCLUSIONS

Patients treated with etanercept, infliximab, or abatacept, had similar odds of achieving DAS28-remission compared to those treated with adalimumab, irrespectively of the BMI category, with the exception of infliximab in female patients. Compared to adalimumab, higher odds of DAS28 remission were observed in overweight female patients treated with infliximab, while, conversely, lower odds were observed in female obese users of infliximab. Additionally, the differential odds of achieving RADAI-5 remission between etanercept and adalimumab in overweight patients requires further attention. Ultimately, while the study findings suggested differential effectiveness of biologics depending on the BMI and sex of the patient, the selection of an optimal biologic in patients with abnormal BMI remains of interest, and the role of infliximab and etanercept depending on BMI may be further investigated.

## Data Availability

Restrictions apply to the availability of these data. Data was obtained from the Swiss Clinical Quality Management in Rheumatic Diseases (SCQM) and its availability requires received approval and permission from the license holder (SCQM).

## Competing interests

EV-Y, TB, and AMB declare no competing interests. A.F. received funding from AbbVie, Bristol Myers Squibb, Galapagos, and Pfizer to the Geneva University Hospital, and consultancies or honoraria from AbbVie, Bristol Myers Squibb, Eli-Lilly, MSD and Pfizer to the Geneva University Hospital.

## Contributorship

EV-Y and AMB contributed to the study conceptualisation and methodology. EV-Y performed data curation, formal analysis, visualisation, and investigation; E.V.-Y. wrote the original draft manuscript. All authors contributed to the interpretations of the findings, provided input, and read and approved the final manuscript.

## Ethical approval for this work

This study was reviewed by the ethics commission of the Canton of Zurich (KEK: Req-2020-00045). Pseudonymized data, without access to the code key, was provided by the Swiss Clinical Quality Management in Rheumatic Diseases (SCQM) registry to the researchers. Therefore, the commission waived the need for a full ethics authorisation.

## Acknowledgements

We thank all patients and clinicians contributing to the SCQM registry, as well as the entire SCQM staff. A list of rheumatology offices and hospitals which contribute to the SCQM registry can be found at http://www.scqm.ch/institutions. A list of financial supporters of SCQM can be found at http://www.scqm.ch/sponsors. AMB acknowledges that her professorship is partly endowed by the Swiss National Pharmacy Association (PharmaSuisse) and the ETH Foundation.

## Funding

Not applicable.

## Notes

### Funding Statement

This study did not receive any funding.

### Author Declarations

This study was reviewed by the ethics commission of the Canton of Zurich (KEK: Req-2020-00045). Pseudonymized data, without access to the code key, was provided by the Swiss Clinical Quality Management in Rheumatic Diseases (SCQM) registry to the researchers. Therefore, the commission waived the need for a full ethics authorization.

