## Supplementary for "Comparative effectiveness of biologics in patients with rheumatoid arthritis stratified by body mass index and sex: a cohort study in SCQM"

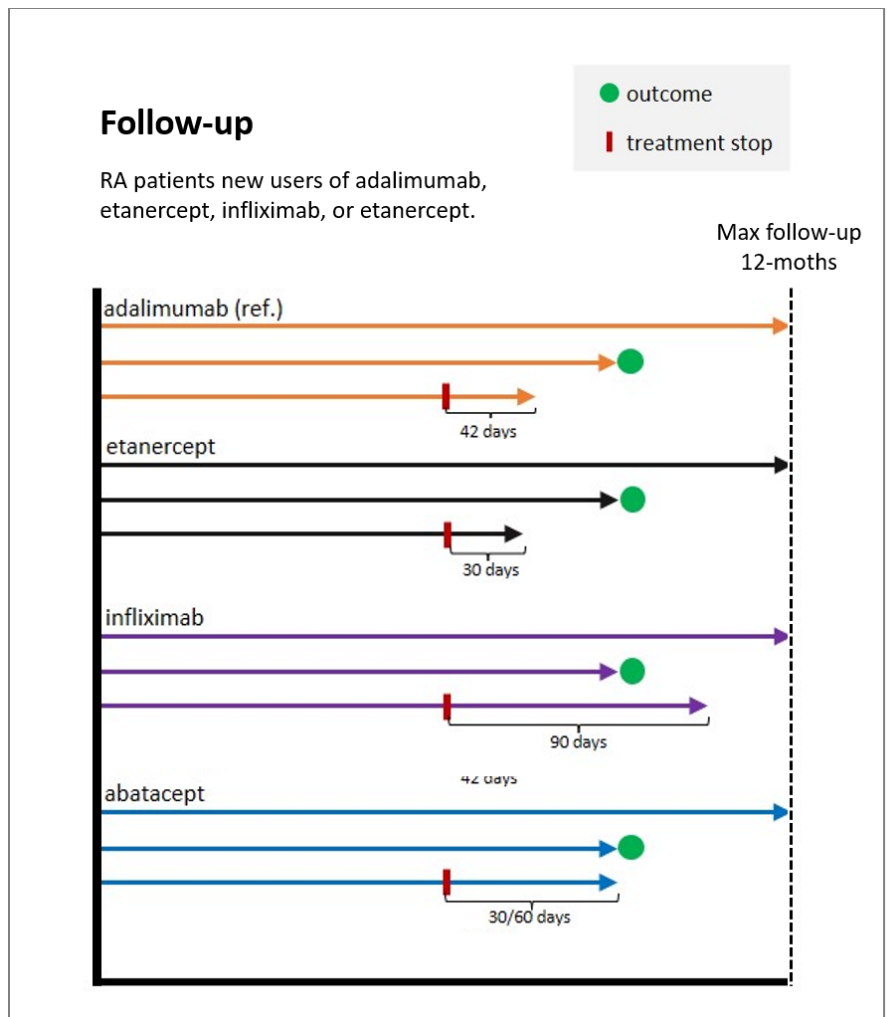

**Supplementary Figure S1** Comparative effectiveness analysis. Abbreviations: RA rheumatoid arthritis; Max maximum; ref. reference.

**Supplementary Table S1** Variables included in the multiple imputation, conducted on outcome and cohort basis.

| Variable | Included for DAS28 | Included for RADAI-5 | Included for individual | Predicted | Predictor | Method | Levels |
| --- | --- | --- | --- | --- | --- | --- | --- |
| **Baseline variables:** |  |  |  |  |  |  |  |
| Biologic agent | yes | yes | yes | - | yes | - | adalimumab; etanercept; infliximab; abatacept. |
| Sex<sup>a</sup> | yes<sup>a</sup> | yes<sup>a</sup> | yes<sup>a</sup> | - | yes<sup>a</sup> | - | female; male. |
| Age | yes | yes | yes | - | yes | - | - |
| BMI kg/m<sup>2</sup> | yes | yes | yes | - | yes | - | - |
| Smoker ever before | yes | yes | yes | - | yes | - | yes; no. |
| Disease duration, years | yes | yes | yes | yes | yes | pmm | - |
| Year of index date | yes | yes | yes | - | yes | - | - |
| csDMARD at index | yes | yes | yes | - | yes | - | yes; no. |
| Prednisone at index | yes | yes | yes | - | yes | - | yes; no. |
| Seropositivity | yes | yes | yes | yes | yes | logreg | yes; no. |
| ESR | yes | yes | yes | yes | yes | pmm | - |
| CRP | - | - | yes | yes | yes | pmm | - |
| SJC28 | yes | yes | yes | yes | yes | pmm | - |
| TJC28 | yes | yes | yes | yes | yes | pmm | - |
| DAS28-ESR | yes | yes | yes | yes | yes<sup>b</sup> | passive imputation<sup>c</sup> | - |
| RADAI-5 | yes | yes | yes | yes | yes | pmm | - |
| HAQ | yes | yes | yes | yes | yes | pmm | - |
| **Follow-up variables:** |  |  |  |  |  |  |  |
| Treatment duration (time to stop or censor) | yes | yes | yes | - | yes | - | - |
| Treatment stop, or censor | yes | yes | yes | - | yes | - | censor; stop. |
| Reason for treatment stop (or switch) | yes | yes | yes | yes | yes | polyreg | adverse event; not effective; remission; other; stop/censor >3years. |
| DAS28-ESR<sub>fu</sub> | yes | - | - | yes | yes | pmm | - |
| RADAI-5<sub>fu</sub> | - | yes | - | yes | yes | pmm | - |
| ESR<sub>fu</sub> | - | - | yes | yes | yes | pmm | - |
| CRP<sub>fu</sub> | - | - | yes | yes | yes | pmm | - |
| TJC28<sub>fu</sub> | - | - | yes | yes | yes | pmm | - |
| SJC28<sub>fu</sub> | - | - | yes | yes | yes | pmm | - |
| Outcome DAS28-remission | yes | - | - | yes | - | passive imputation [ESR<sub>fu</sub><2.6 → yes] | yes; no. |
| Outcome DAS28-LDA | yes | - | - | yes | - | passive imputation [ESR<sub>fu</sub><3.2 → yes] | yes; no. |
| Outcome RADAI-5 | - | yes | - | yes | - | passive imputation [RADAI<sub>fu</sub>≤1.4 → yes] | yes; no. |
| Outcome ΔESR | - | - | yes | yes | - | passive imputation [ESR<sub>fu</sub> - ESR<sub>baseline</sub>] | - |
| Outcome ΔCPR | - | - | yes | yes | - | passive imputation [CRP<sub>fu</sub> - CRP<sub>baseline</sub>] | - |
| Outcome ΔTJC28 | - | - | yes | yes | - | passive imputation [TJC28<sub>fu</sub> - TJC28<sub>baseline</sub>] | - |
| Outcome ΔSJC28 | - | - | yes | yes | - | passive imputation [SJC28<sub>fu</sub> - SJC28<sub>baseline</sub>] | - |

When unspecified, variables correspond to baseline data. Otherwise, it is specified as fu (follow-up) or outcome.

<sup>a</sup> Sex not included as variable in the sex-stratified cohorts.

<sup>b</sup> Baseline DAS28 not used as predictor for baseline ESR, TJC28, and SJC28.

<sup>c</sup> Baseline DAS28 passive imputation: $DAS28ESR = (0.56 \times \sqrt{tjc28} + 0.28 \times \sqrt{sjc28} + 0.7 \times \ln(ESR)) \times 1.08 + 0.16$

Abbreviations: DAS28 28-joint Disease Activity Score; RADAI-5 Rheumatoid Arthritis Disease Activity Index-Five; BMI body mass index; csDMARD conventional synthetic disease modifying anti-rheumatic drug; ESR erythrocyte sedimentation rate; fu follow-up; logreg logistic regression; pmm predictive mean matching.

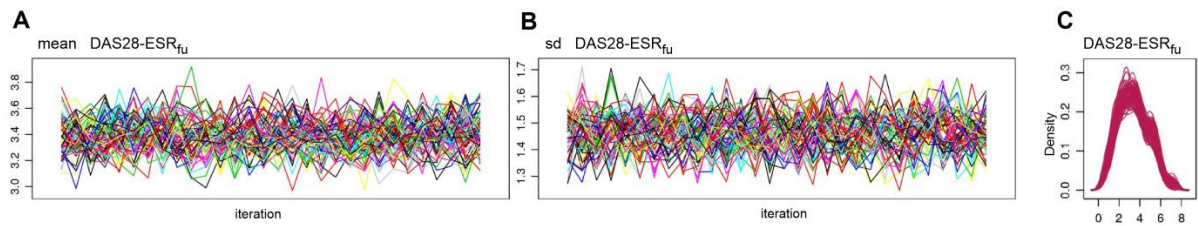

**Supplementary Figure S2** Visualization of the coverage of the multiple imputation by chain equations (MICE) or confounder-adjusted response rate with attrition correction (CARRAC) of DAS28-ESR for the outcome DAS28-remission in the obese cohort with a maximum follow-up of 12-months, particularly the variation of the mean (A) and standard deviation (B) of the DAS28-ESR at follow-up. On the right (C), density plot depicting the distribution of the DAS28-ESR at follow-up. In C, the distribution in the original data set is depicted with a blue line, and distribution in each imputed dataset is depicted with a red line per dataset (Blue line not visible, covered by the red lines). Abbreviations: sd standard deviation; DAS28 28-joint Disease Activity Score; ESR erythrocyte sedimentation rate; fu follow-up.

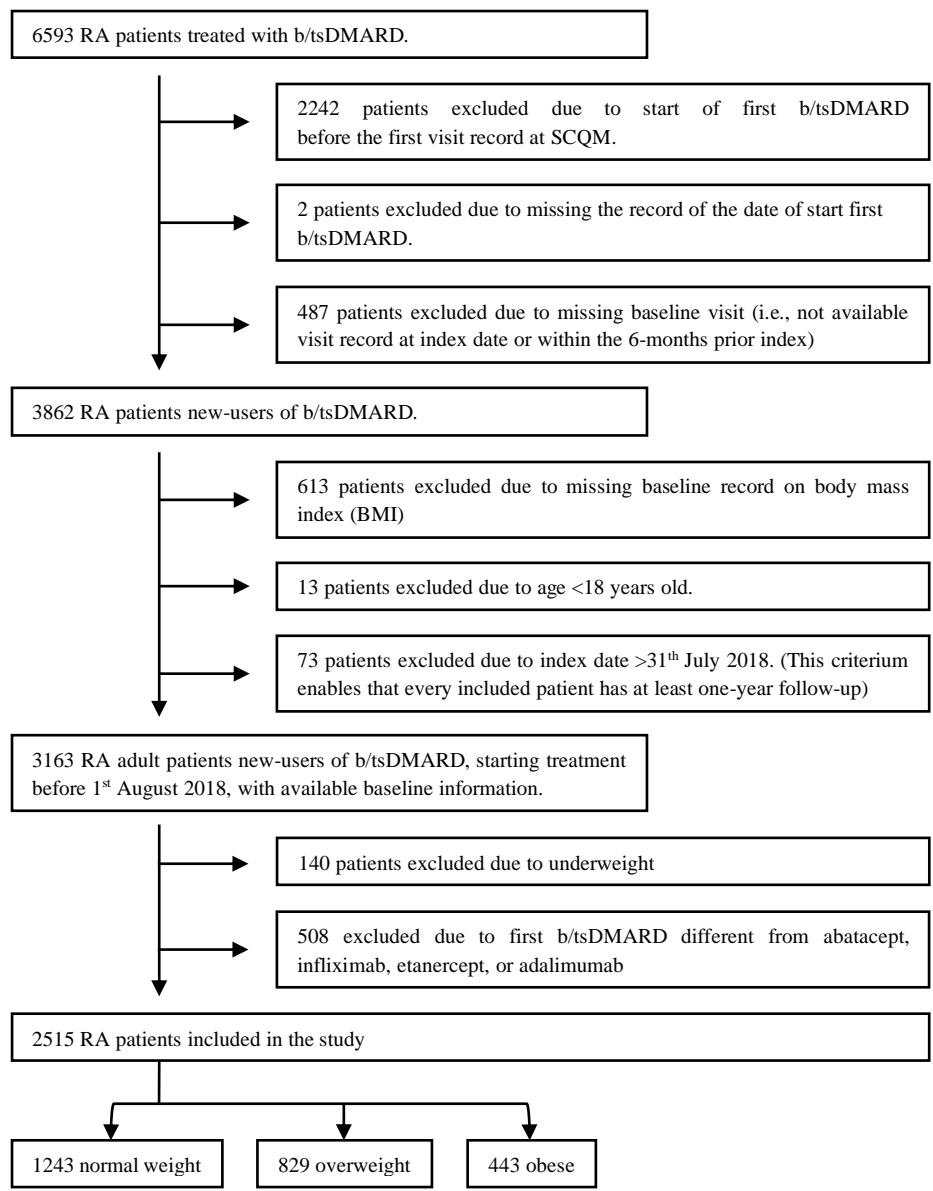

**Supplementary Figure S3** Flow chart of the inclusion and exclusion criteria. Abbreviations: RA rheumatoid arthritis; b/tsDMARD biologic or targeted synthetic disease modifying anti-rheumatic drug; SCQM Swiss Quality Management of Rheumatic Diseases; BMI body mass index.

**Supplementary Table S3** Obese cohort, patient characteristics at baseline, stratified by first b/tsDMARD adalimumab, etanercept, infliximab, and abatacept.

| Obese | Adalimumab | Etanercept |  | Infliximab |  | Abatacept |  |
| --- | --- | --- | --- | --- | --- | --- | --- |
| n | 178 | 150 | p value | 73 | p value | 42 | p value |
| Women (%) | 130 (73.0) | 124 (82.7) | 0.052 | 56 (76.7) | 0.656 | 27 (64.3) | 0.348 |
| Age index (mean (SD)) | 56.60 (11.99) | 56.98 (11.63) | 0.777 | 55.86 (10.77) | 0.644 | 59.27 (10.00) | 0.183 |
| RA duration, years (mean (SD)) | 6.72 (8.67) | 7.75 (8.45) | 0.285 | 7.92 (8.61) | 0.331 | 5.80 (7.42) | 0.531 |
| missing/unknown | 5 (2.8) | 7 (4.7) |  | 4 (5.5) |  | 0 (0) |  |
| Year of index date (mean (SD)) | 2007.85 (3.79) | 2007.53 (4.56) | 0.487 | 2005.68 (4.02) | <0.001 | 2013.19 (2.52) | <0.001 |
| BMI kg/m2 (mean (SD)) | 33.90 (3.86) | 33.79 (3.85) | 0.794 | 33.65 (3.09) | 0.621 | 33.91 (3.99) | 0.987 |
| High educational level (%) | 11 (6.2) | 4 (2.7) | 0.268 | 4 (5.5) | 1.000 | 4 (9.5) | 0.690 |
| missing/unknown | 58 (32.6) | 57 (38.0) |  | 27 (37.0) |  | 13 (31.0) |  |
| Ever smoker (%) | 49 (27.5) | 41 (27.3) | 1.000 | 11 (15.1) | 0.053 | 22 (52.4) | 0.004 |
| csDMARD at index date (%) | 134 (75.3) | 96 (64.0) | 0.036 | 60 (82.2) | 0.307 | 33 (78.6) | 0.804 |
| Prednisone at index date (%) | 66 (37.1) | 60 (40.0) | 0.669 | 41 (56.2) | 0.008 | 18 (42.9) | 0.605 |
| Seropositive (%) | 129 (72.5) | 102 (68.0) | 0.691 | 56 (76.7) | 0.537 | 32 (76.2) | 0.765 |
| missing/unknown | 20 (11.2) | 21 (14.0) |  | 8 (11.0) |  | 1 (2.4) |  |
| ESR (mean (SD)) | 24.22 (18.46) | 23.73 (16.09) | 0.809 | 29.43 (18.87) | 0.053 | 22.00 (18.05) | 0.518 |
| missing/unknown | 14 (7.9) | 18 (12.0) |  | 5 (6.8) |  | 7 (16.7) |  |
| CRP (mean (SD)) | 1.44 (1.20) | 1.37 (1.42) | 0.765 | 1.38 (1.08) | 0.842 | 1.06 (0.97) | 0.106 |
| missing/unknown | 105 (59.0) | 88 (58.7) |  | 54 (74.0) |  | 6 (14.3) |  |
| Tender joint counts 28 (mean (SD)) | 7.25 (6.92) | 7.86 (6.75) | 0.432 | 8.14 (7.20) | 0.367 | 7.19 (6.66) | 0.963 |
| missing/unknown | 8 (4.5) | 11 (7.3) |  | 2 (2.7) |  | 5 (11.9) |  |
| Swollen joint counts 28 (mean (SD)) | 6.75 (5.72) | 6.63 (5.79) | 0.865 | 7.38 (6.37) | 0.449 | 5.73 (4.85) | 0.317 |
| missing/unknown | 9 (5.1) | 11 (7.3) |  | 2 (2.7) |  | 5 (11.9) |  |
| Physician global disease activity (mean (SD)) | 4.94 (1.85) | 4.93 (1.95) | 0.950 | 5.29 (2.13) | 0.344 | 4.09 (1.99) | 0.024 |
| missing/unknown | 70 (39.33) | 68 (45.33) |  | 35 (47.95) |  | 9 (21.43) |  |
| DAS28-ESR (mean (SD)) | 4.40 (1.42) | 4.54 (1.32) | 0.406 | 4.72 (1.40) | 0.118 | 4.18 (1.46) | 0.399 |
| missing/unknown | 15 (8.43) | 18 (12) |  | 5 (6.85) |  | 8 (19.05) |  |
| DAS28-CRP (mean (SD)) | 4.45 (1.15) | 4.36 (1.08) | 0.648 | 3.78 (1.10) | 0.026 | 4.06 (1.22) | 0.113 |
| missing/unknown | 105 (58.99) | 88 (58.67) |  | 54 (73.97) |  | 7 (16.67) |  |
| RADAI-5 (mean (SD)) | 4.91 (2.04) | 5.33 (2.18) | 0.107 | 5.15 (2.12) | 0.425 | 4.35 (2.36) | 0.209 |
| missing/unknown | 28 (15.73) | 35 (23.33) |  | 8 (10.96) |  | 16 (38.1) |  |
| HAQ (mean (SD)) | 1.21 (0.74) | 1.32 (0.73) | 0.235 | 1.38 (0.77) | 0.115 | 0.95 (0.70) | 0.093 |
| missing/unknown | 23 (12.92) | 30 (20) |  | 6 (8.22) |  | 15 (35.71) |  |
| Osteoporosis | 24 (13.5) | 23 (15.3) | 0.750 | 11 (15.1) | 0.898 | 4 (9.5) | 0.663 |
| Fibromyalgia | 2 (1.1) | 8 (5.3) | 0.059 | 0 (0.0) | 0.898 | 1 (2.4) | 1.000 |
| Other rheumatological disease | 61 (34.3) | 62 (41.3) | 0.229 | 25 (34.2) | 1.000 | 8 (19.0) | 0.084 |
| Psoriasis | 2 (1.1) | 2 (1.3) | 1.000 | 1 (1.4) | 1.000 | 0 (0.0) | 1.000 |
| Hyperlipidemia | 15 (8.4) | 11 (7.3) | 0.873 | 5 (6.8) | 0.871 | 10 (23.8) | 0.011 |
| Cardiac/cardiovascular event/disease | 84 (47.2) | 84 (56.0) | 0.139 | 32 (43.8) | 0.730 | 30 (71.4) | 0.008 |
| Cancer | 7 (3.9) | 4 (2.7) | 0.744 | 1 (1.4) | 0.513 | 4 (9.5) | 0.270 |
| Depression/anxiety | 25 (14.0) | 31 (20.7) | 0.150 | 12 (16.4) | 0.772 | 7 (16.7) | 0.849 |
| Diabetes | 17 (9.6) | 23 (15.3) | 0.154 | 9 (12.3) | 0.669 | 8 (19.0) | 0.140 |
| Fractures, surgeries, musculoskeletal system | 15 (8.4) | 8 (5.3) | 0.381 | 7 (9.6) | 0.960 | 3 (7.1) | 1.000 |

Values are the number and column percentage, unless otherwise specified. Significance tests compare each drug of interest to Adalimumab, using chi-squared test for categorical variables, and t-test for continuous variables. For these tests, missing values did not function as a grouping variable.

Abbreviations: SD standard deviation; RA rheumatoid arthritis; BMI body mass index; csDMARD conventional synthetic disease modifying anti-rheumatic drug; ESR erythrocyte sedimentation rate; CRP C-reactive protein; DAS28-ESR 28-joint Disease Activity Score using erythrocyte sedimentation rate; DAS28-CRP 28-joint Disease Activity Score using C-reactive protein; RADAI-5 Rheumatoid Arthritis Disease Activity Index-Five; HAQ Health Assessment Questionnaire.

Additional information on seropositivity and comorbidities: Seropositivity was calculated using both rheumatoid factor (RF) and anti-cyclic citrullinated peptide (anti-CCP) antibodies. Osteoporosis includes osteoporosis record or medication with bisphosphonates, denosumab, or teriparatide; Other rheumatological disease includes gout, lupus, osteoarthritis, Sjogren syndrome, degenerative spine disease, degenerative spondylopathy, other connective tissue disease, other rheumatological disease; Cardiac/cardiovascular event/disease includes myocardial infarction, heart infarct, heart failure, heart insufficiency, cardiac insufficiency, coronary heart disease, coronary cardiac disease, heart problem, heart disease, angina pectoris, rhythm disorder, artery intervention, stroke transient ischemic attack, cerebrovascular disease, deep venous thrombosis, peripheral vascular disease, pulmonary embolism, blood thinners, hypertension, hypotension, other cardiovascular disease, and medication with platelet aggregation inhibitors, antihypertensives, or statins; Depression/anxiety includes record of the disease or medication with antidepressants.

**Supplementary Table S4** Overweight cohort, patient characteristics at baseline, stratified by first b/tsDMARD adalimumab, etanercept, infliximab, and abatacept.

| Overweight | Adalimumab | Etanercept |  | Infliximab |  | Abatacept |  |
| --- | --- | --- | --- | --- | --- | --- | --- |
| n | 336 | 296 | p value | 150 | p value | 47 | p value |
| Women (%) | 215 (64.0) | 203 (68.6) | 0.257 | 91 (60.7) | 0.549 | 30 (63.8) | 1.000 |
| Age index (mean (SD)) | 57.28 (12.52) | 57.81 (12.32) | 0.589 | 56.87 (11.35) | 0.734 | 62.99 (10.81) | 0.003 |
| RA duration, years (mean (SD)) | 7.10 (7.76) | 8.45 (9.21) | 0.050 | 7.66 (8.08) | 0.477 | 6.60 (8.45) | 0.690 |
| missing/unknown | 12 (3.57) | 7 (2.36) |  | 4 (2.67) |  | 3 (6.38) |  |
| Year of index date (mean (SD)) | 2007.68 (3.56) | 2006.72 (5.02) | 0.005 | 2005.93 (4.69) | <0.001 | 2012.32 (2.60) | <0.001 |
| BMI kg/m<sup>2</sup> (mean (SD)) | 27.16 (1.35) | 27.21 (1.33) | 0.643 | 27.13 (1.37) | 0.829 | 27.19 (1.41) | 0.885 |
| High educational level (%) | 29 (8.6) | 21 (7.1) | 0.833 | 12 (8.0) | 0.892 | 6 (12.8) | 0.629 |
| missing/unknown | 98 (29.2) | 106 (35.8) |  | 41 (27.3) |  | 11 (23.4) |  |
| Ever smoker (%) | 94 (28.0) | 76 (25.7) | 0.575 | 43 (28.7) | 0.962 | 22 (46.8) | 0.014 |
| csDMARD at index date (%) | 226 (67.3) | 189 (63.9) | 0.414 | 126 (84.0) | <0.001 | 34 (72.3) | 0.595 |
| Prednisone at index date (%) | 134 (39.9) | 133 (44.9) | 0.229 | 84 (56.0) | 0.001 | 13 (27.7) | 0.146 |
| Seropositive (%) | 244 (72.6) | 211 (71.3) | 0.679 | 119 (79.3) | 0.731 | 37 (78.7) | 0.697 |
| missing/unknown | 47 (14.0) | 41 (13.9) |  | 12 (8.0) |  | 5 (10.6) |  |
| ESR (mean (SD)) | 24.82 (19.57) | 25.87 (22.59) | 0.555 | 25.44 (21.41) | 0.766 | 28.92 (18.07) | 0.221 |
| missing/unknown | 39 (11.61) | 21 (7.09) |  | 9 (6) |  | 9 (19.15) |  |
| CRP (mean (SD)) | 2.12 (3.25) | 1.71 (3.10) | 0.315 | 1.18 (1.68) | 0.057 | 1.30 (1.48) | 0.121 |
| missing/unknown | 208 (61.9) | 181 (61.15) |  | 101 (67.33) |  | 6 (12.77) |  |
| Tender joint counts 28 (mean (SD)) | 7.55 (6.90) | 7.95 (7.69) | 0.507 | 7.14 (6.85) | 0.552 | 6.00 (5.88) | 0.161 |
| missing/unknown | 33 (9.82) | 12 (4.05) |  | 5 (3.33) |  | 4 (8.51) |  |
| Swollen joint counts 28 (mean (SD)) | 6.90 (5.51) | 6.68 (5.93) | 0.639 | 7.92 (5.98) | 0.077 | 5.82 (5.18) | 0.219 |
| missing/unknown | 33 (9.82) | 12 (4.05) |  | 5 (3.33) |  | 3 (6.38) |  |
| Physician global disease activity (mean (SD)) | 4.92 (2.27) | 4.90 (2.14) | 0.907 | 5.56 (2.04) | 0.033 | 4.28 (1.89) | 0.091 |
| missing/unknown | 136 (40.48) | 140 (47.3) |  | 73 (48.67) |  | 7 (14.89) |  |
| DAS28-ESR (mean (SD)) | 4.47 (1.34) | 4.42 (1.50) | 0.690 | 4.42 (1.48) | 0.747 | 4.49 (1.15) | 0.933 |
| missing/unknown | 41 (12.2) | 21 (7.09) |  | 9 (6) |  | 9 (19.15) |  |
| DAS28-CRP (mean (SD)) | 4.28 (1.20) | 4.19 (1.31) | 0.584 | 4.01 (1.27) | 0.189 | 3.98 (1.07) | 0.161 |
| missing/unknown | 209 (62.2) | 181 (61.15) |  | 101 (67.33) |  | 7 (14.89) |  |
| RADAI-5 (mean (SD)) | 4.84 (2.08) | 5.01 (2.17) | 0.358 | 4.98 (2.18) | 0.528 | 4.87 (2.35) | 0.935 |
| missing/unknown | 73 (21.7) | 44 (14.9) |  | 24 (16) |  | 16 (34) |  |
| HAQ (mean (SD)) | 1.08 (0.70) | 1.13 (0.74) | 0.408 | 1.24 (0.72) | 0.030 | 0.94 (0.69) | 0.272 |
| missing/unknown | 63 (18.75) | 38 (12.84) |  | 14 (9.33) |  | 14 (29.79) |  |
| Osteoporosis | 59 (17.6) | 61 (20.6) | 0.382 | 33 (22.0) | 0.303 | 11 (23.4) | 0.442 |
| Fibromyalgia | 0 (0.0) | 9 (3.0) | 0.004 | 5 (3.3) | 0.004 | 2 (4.3) | 0.007 |
| Other rheumatological disease | 101 (30.1) | 97 (32.8) | 0.517 | 45 (30.0) | 1.000 | 13 (27.7) | 0.868 |
| Psoriasis | 2 (0.6) | 3 (1.0) | 0.887 | 0 (0.0) | 0.857 | 1 (2.1) | 0.816 |
| Hyperlipidemia | 16 (4.8) | 25 (8.4) | 0.086 | 8 (5.3) | 0.967 | 7 (14.9) | 0.016 |
| Cardiac/cardiovascular event/disease | 123 (36.6) | 117 (39.5) | 0.501 | 58 (38.7) | 0.740 | 21 (44.7) | 0.363 |
| Cancer | 6 (1.8) | 13 (4.4) | 0.093 | 1 (0.7) | 0.586 | 2 (4.3) | 0.572 |
| Depression/anxiety | 31 (9.2) | 42 (14.2) | 0.068 | 27 (18.0) | 0.009 | 2 (4.3) | 0.390 |
| Diabetes | 26 (7.7) | 19 (6.4) | 0.625 | 8 (5.3) | 0.443 | 8 (17.0) | 0.068 |
| Fractures, surgeries, musculoskeletal system | 26 (7.7) | 34 (11.5) | 0.142 | 13 (8.7) | 0.867 | 2 (4.3) | 0.575 |

Values are the number and column percentage, unless otherwise specified. Significance tests compare each drug of interest to Adalimumab, using chi-squared test for categorical variables, and t-test for continuous variables. For these tests, missing values did not function as a grouping variable.

Abbreviations: SD standard deviation; RA rheumatoid arthritis; BMI body mass index; csDMARD conventional synthetic disease modifying anti-rheumatic drug; ESR erythrocyte sedimentation rate; CRP C-reactive protein; DAS28-ESR 28-joint Disease Activity Score using erythrocyte sedimentation rate; DAS28-CRP 28-joint Disease Activity Score using C-reactive protein; RADAI-5 Rheumatoid Arthritis Disease Activity Index-Five; HAQ Health Assessment Questionnaire.

For additional information on seropositivity and comorbidities see footnote in Supplementary Table S3.

**Supplementary Table S5** Normal weight cohort, patient characteristics at baseline, stratified by first b/tsDMARD adalimumab, etanercept, infliximab, and abatacept.

| Normal weight | Adalimumab<br>442 | Etanercept<br>482 | p value | Infliximab<br>259 | p value | Abatacept<br>60 | p value |
| --- | --- | --- | --- | --- | --- | --- | --- |
| Women (%) | 365 (82.6) | 393 (81.5) | 0.744 | 207 (79.9) | 0.438 | 51 (85.0) | 0.776 |
| Age index (mean (SD)) | 53.24 (13.78) | 54.32 (14.68) | 0.253 | 53.15 (14.28) | 0.936 | 61.97 (14.34) | <0.001 |
| RA duration, years (mean (SD)) | 8.29 (9.16) | 8.66 (8.96) | 0.537 | 9.82 (9.32) | 0.036 | 9.15 (10.51) | 0.506 |
| missing/unknown | 10 (2.26) | 13 (2.7) |  | 6 (2.32) |  | 2 (3.33) |  |
| Year of index date (mean (SD)) | 2007.33 (3.46) | 2005.82 (4.90) | <0.001 | 2004.81 (4.04) | <0.001 | 2012.72 (2.68) | <0.001 |
| BMI kg/m2 (mean (SD)) | 22.17 (1.77) | 22.07 (1.82) | 0.413 | 22.17 (1.82) | 0.973 | 22.43 (1.74) | 0.278 |
| High educational level (%) | 50 (11.3) | 47 (9.8) | 0.630 | 24 (9.3) | 0.702 | 6 (10.0) | 0.579 |
| missing/unknown | 140 (31.7) | 165 (34.2) |  | 96 (37.1) |  | 11 (18.3) |  |
| Ever smoker (%) | 122 (27.6) | 119 (24.7) | 0.351 | 55 (21.2) | 0.075 | 18 (30.0) | 0.814 |
| csDMARD at index date (%) | 316 (71.5) | 265 (55.0) | <0.001 | 208 (80.3) | 0.012 | 38 (63.3) | 0.250 |
| Prednisone at index date (%) | 168 (38.0) | 193 (40.0) | 0.572 | 124 (47.9) | 0.013 | 27 (45.0) | 0.367 |
| Seropositive (%) | 325 (73.5) | 379 (78.6) | 0.175 | 208 (80.3) | 0.012 | 46 (76.7) | 0.456 |
| missing/unknown | 56 (12.7) | 50 (10.4) |  | 32 (12.4) |  | 2 (3.3) |  |
| ESR (mean (SD)) | 23.49 (20.10) | 25.78 (23.17) | 0.134 | 26.11 (24.61) | 0.149 | 26.87 (22.18) | 0.288 |
| missing/unknown | 58 (13.12) | 43 (8.92) |  | 24 (9.27) |  | 14 (23.33) |  |
| CRP (mean (SD)) | 1.28 (1.57) | 1.32 (1.61) | 0.855 | 1.44 (1.98) | 0.561 | 1.47 (1.92) | 0.506 |
| missing/unknown | 299 (67.65) | 350 (72.61) |  | 200 (77.22) |  | 13 (21.67) |  |
| Tender joint counts 28 (mean (SD)) | 6.70 (6.41) | 6.69 (6.66) | 0.989 | 6.59 (6.75) | 0.837 | 6.44 (5.59) | 0.783 |
| missing/unknown | 40 (9.05) | 26 (5.39) |  | 18 (6.95) |  | 8 (13.33) |  |
| Swollen joint counts 28 (mean (SD)) | 6.56 (5.67) | 6.83 (6.09) | 0.506 | 8.42 (6.92) | <0.001 | 6.17 (5.31) | 0.635 |
| missing/unknown | 39 (8.82) | 26 (5.39) |  | 16 (6.18) |  | 7 (11.67) |  |
| Physician GDA (mean (SD)) | 4.98 (2.12) | 5.14 (2.17) | 0.397 | 5.23 (2.18) | 0.269 | 4.40 (1.75) | 0.101 |
| missing/unknown | 200 (45.25) | 230 (47.72) |  | 131 (50.58) |  | 18 (30) |  |
| DAS28-ESR (mean (SD)) | 4.29 (1.40) | 4.30 (1.43) | 0.926 | 4.35 (1.57) | 0.619 | 4.33 (1.18) | 0.836 |
| missing/unknown | 60 (13.57) | 44 (9.13) |  | 26 (10.04) |  | 15 (25) |  |
| DAS28-CRP (mean (SD)) | 4.11 (1.22) | 4.14 (1.13) | 0.827 | 3.98 (1.19) | 0.483 | 4.02 (1.18) | 0.650 |
| missing/unknown | 301 (68.1) | 351 (72.82) |  | 202 (77.99) |  | 14 (23.33) |  |
| RADAI-5 (mean (SD)) | 4.61 (2.22) | 4.69 (2.14) | 0.61 | 4.59 (2.10) | 0.888 | 4.34 (1.81) | 0.434 |
| missing/unknown | 88 (19.9) | 63 (13.1) |  | 45 (17.4) |  | 16 (26.7) |  |
| HAQ (mean (SD)) | 0.96 (0.70) | 1.03 (0.72) | 0.173 | 1.12 (0.72) | 0.010 | 0.78 (0.64) | 0.089 |
| missing/unknown | 80 (18.1) | 59 (12.24) |  | 29 (11.2) |  | 14 (23.33) |  |
| Osteoporosis | 88 (19.9) | 101 (21.0) | 0.755 | 61 (23.6) | 0.297 | 23 (38.3) | 0.002 |
| Fibromyalgia | 1 (0.2) | 1 (0.2) | 1.000 | 4 (1.5) | 0.124 | 0 (0.0) | 1.000 |
| Other rheumatological disease | 99 (22.4) | 118 (24.5) | 0.504 | 70 (27.0) | 0.197 | 16 (26.7) | 0.566 |
| Psoriasis | 5 (1.1) | 2 (0.4) | 0.382 | 2 (0.8) | 0.946 | 0 (0.0) | 0.892 |
| Hyperlipidemia | 9 (2.0) | 14 (2.9) | 0.525 | 3 (1.2) | 0.573 | 7 (11.7) | <0.001 |
| Cardiac/cardiovascular event/disease | 78 (17.6) | 115 (23.9) | 0.025 | 56 (21.6) | 0.233 | 31 (51.7) | <0.001 |
| Cancer | 4 (0.9) | 9 (1.9) | 0.337 | 6 (2.3) | 0.234 | 4 (6.7) | 0.005 |
| Depression/anxiety | 30 (6.8) | 46 (9.5) | 0.160 | 30 (11.6) | 0.040 | 6 (10.0) | 0.523 |
| Diabetes | 6 (1.4) | 21 (4.4) | 0.012 | 4 (1.5) | 1.000 | 4 (6.7) | 0.023 |
| Fractures, surgeries, musculoskeletal system | 43 (9.7) | 60 (12.4) | 0.227 | 38 (14.7) | 0.064 | 3 (5.0) | 0.341 |

Values are the number and column percentage, unless otherwise specified. Significance tests compare each drug of interest to Adalimumab, using chi-squared test for categorical variables, and t-test for continuous variables. For these tests, missing values did not function as a grouping variable.

Abbreviations: SD standard deviation; RA rheumatoid arthritis; BMI body mass index; csDMARD conventional synthetic disease modifying anti-rheumatic drug; ESR erythrocyte sedimentation rate; CRP C-reactive protein; DAS28-ESR 28-joint Disease Activity Score using erythrocyte sedimentation rate; DAS28-CRP 28-joint Disease Activity Score using C-reactive protein; RADAI-5 Rheumatoid Arthritis Disease Activity Index-Five; HAQ Health Assessment Questionnaire.

For additional information on seropositivity and comorbidities see footnote in Supplementary Table S3.

**Supplementary Table S6** Obese female cohort, patient characteristics at baseline, stratified by first b/tsDMARD adalimumab, etanercept, infliximab, and abatacept.

| Obese female | adalimumab<br>130 | etanercept<br>124 | p value | infliximab<br>56 | p value | abatacept<br>27 | p value |
| --- | --- | --- | --- | --- | --- | --- | --- |
| Age index (mean (SD)) | 56.70 (12.45) | 56.27 (11.65) | 0.778 | 55.87 (11.33) | 0.669 | 60.33 (10.55) | 0.159 |
| RA duration, years (mean (SD)) | 6.92 (8.60) | 7.19 (7.66) | 0.798 | 7.97 (8.91) | 0.460 | 6.33 (7.94) | 0.744 |
| missing/unknown | 5 (3.85) | 5 (4.03) |  | 2 (3.57) |  | 0 (0) |  |
| Year of index date (mean (SD)) | 2007.86 (3.80) | 2008.00 (4.52) | 0.791 | 2006.16 (4.12) | 0.007 | 2013.63 (2.20) | <0.001 |
| BMI kg/m<sup>2</sup> (mean (SD)) | 34.25 (4.13) | 34.12 (4.07) | 0.792 | 33.71 (3.10) | 0.376 | 34.69 (4.65) | 0.626 |
| High educational level (%) | 10 (7.7) | 3 (2.4) | 0.187 | 3 (5.4) | 1.000 | 3 (11.1) | 0.804 |
| missing/unknown | 36 (27.7) | 49 (39.5) |  | 23 (41.1) |  | 8 (29.6) |  |
| Ever smoker (%) | 34 (26.2) | 31 (25.0) | 0.947 | 8 (14.3) | 0.113 | 12 (44.4) | 0.095 |
| csDMARD at index date (%) | 95 (73.1) | 83 (66.9) | 0.352 | 45 (80.4) | 0.384 | 21 (77.8) | 0.791 |
| Prednisone at index date (%) | 42 (32.3) | 51 (41.1) | 0.184 | 29 (51.8) | 0.019 | 12 (44.4) | 0.324 |
| Seropositive (%) | 94 (72.3) | 84 (67.7) | 0.408 | 44 (78.6) | 0.620 | 18 (66.7) | 0.247 |
| missing/unknown | 15 (11.5) | 14 (11.3) |  | 5 (8.9) |  | 1 (3.7) |  |
| ESR (mean (SD)) | 25.50 (18.54) | 24.62 (16.21) | 0.704 | 30.59 (17.45) | 0.097 | 19.79 (14.28) | 0.156 |
| missing/unknown | 11 (8.46) | 17 (13.71) |  | 5 (8.93) |  | 3 (11.11) |  |
| CRP (mean (SD)) | 1.32 (1.11) | 1.30 (1.29) | 0.931 | 1.35 (1.04) | 0.922 | 0.96 (0.86) | 0.169 |
| missing/unknown | 75 (57.69) | 70 (56.45) |  | 39 (69.64) |  | 3 (11.11) |  |
| Tender joint counts 28 (mean (SD)) | 7.38 (6.64) | 7.91 (6.70) | 0.540 | 8.19 (7.39) | 0.473 | 5.54 (5.12) | 0.202 |
| missing/unknown | 6 (4.62) | 11 (8.87) |  | 2 (3.57) |  | 3 (11.11) |  |
| Swollen joint counts 28 (mean (SD)) | 6.82 (5.37) | 6.48 (5.69) | 0.634 | 7.06 (6.59) | 0.804 | 4.08 (3.62) | 0.018 |
| missing/unknown | 7 (5.38) | 11 (8.87) |  | 2 (3.57) |  | 3 (11.11) |  |
| Physician GDA (mean (SD)) | 5.11 (1.92) | 4.96 (1.89) | 0.624 | 5.19 (2.11) | 0.863 | 3.43 (1.60) | <0.001 |
| missing/unknown | 48 (36.92) | 53 (42.74) |  | 29 (51.79) |  | 6 (22.22) |  |
| DAS28-ESR (mean (SD)) | 4.54 (1.31) | 4.58 (1.27) | 0.819 | 4.78 (1.36) | 0.284 | 3.95 (1.26) | 0.048 |
| missing/unknown | 12 (9.23) | 17 (13.71) |  | 5 (8.93) |  | 4 (14.81) |  |
| DAS28-CRP (mean (SD)) | 4.45 (1.09) | 4.29 (1.08) | 0.458 | 3.81 (1.16) | 0.041 | 3.79 (1.13) | 0.019 |
| missing/unknown | 75 (57.69) | 70 (56.45) |  | 39 (69.64) |  | 4 (14.81) |  |
| RADAI-5 (mean (SD)) | 5.02 (1.99) | 5.46 (2.27) | 0.146 | 5.06 (2.15) | 0.913 | 4.44 (2.27) | 0.270 |
| missing/unknown | 22 (16.9) | 32 (25.8) |  | 7 (12.5) |  | 9 (33.3) |  |
| HAQ (mean (SD)) | 1.25 (0.77) | 1.36 (0.73) | 0.329 | 1.40 (0.77) | 0.271 | 1.10 (0.65) | 0.408 |
| missing/unknown | 17 (13.08) | 28 (22.58) |  | 6 (10.71) |  | 8 (29.63) |  |
| Osteoporosis | 20 (15.4) | 20 (16.1) | 1.000 | 10 (17.9) | 0.839 | 3 (11.1) | 0.785 |
| Fibromyalgia | 2 (1.5) | 8 (6.5) | 0.091 | 0 (0.0) | 0.874 | 1 (3.7) | 1.000 |
| Other rheumatological disease | 44 (33.8) | 48 (38.7) | 0.499 | 16 (28.6) | 0.593 | 5 (18.5) | 0.182 |
| Psoriasis | 2 (1.5) | 1 (0.8) | 1.000 | 1 (1.8) | 1.000 | 0 (0.0) | 1.000 |
| Hyperlipidemia | 10 (7.7) | 9 (7.3) | 1.000 | 4 (7.1) | 1.000 | 4 (14.8) | 0.418 |
| Cardiac/cardiovascular event/disease | 63 (48.5) | 69 (55.6) | 0.308 | 24 (42.9) | 0.587 | 19 (70.4) | 0.063 |
| Cancer | 6 (4.6) | 3 (2.4) | 0.544 | 1 (1.8) | 0.610 | 2 (7.4) | 0.905 |
| Depression/anxiety | 21 (16.2) | 28 (22.6) | 0.255 | 9 (16.1) | 1.000 | 4 (14.8) | 1.000 |
| Diabetes | 11 (8.5) | 18 (14.5) | 0.187 | 8 (14.3) | 0.348 | 4 (14.8) | 0.508 |
| Fractures, surgeries,<br>musculoskeletal system | 12 (9.2) | 7 (5.6) | 0.397 | 5 (8.9) | 1.000 | 1 (3.7) | 0.572 |

Values are the number and column percentage, unless otherwise specified. Significance tests compare each drug of interest to Adalimumab, using chi-squared test for categorical variables, and t-test for continuous variables. For these tests, missing values did not function as a grouping variable.

Abbreviations: SD standard deviation; RA rheumatoid arthritis; BMI body mass index; csDMARD conventional synthetic disease modifying anti-rheumatic drug; ESR erythrocyte sedimentation rate; CRP C-reactive protein; DAS28-ESR 28-joint Disease Activity Score using erythrocyte sedimentation rate; DAS28-CRP 28-joint Disease Activity Score using C-reactive protein; RADAI-5 Rheumatoid Arthritis Disease Activity Index-Five; HAQ Health Assessment Questionnaire.

For additional information on seropositivity and comorbidities see footnote in Supplementary Table S3.

**Supplementary Table S7** Obese male cohort, patient characteristics at baseline, stratified by first b/tsDMARD adalimumab, etanercept, infliximab, and abatacept.

| Obese male | adalimumab<br>48 | etanercept<br>26 | p value | infliximab<br>17 | p value | abatacept<br>15 | p value |
| --- | --- | --- | --- | --- | --- | --- | --- |
| Age index (mean (SD)) | 56.36 (10.77) | 60.36 (11.12) | 0.136 | 55.82 (8.97) | 0.855 | 57.37 (8.94) | 0.743 |
| RA duration, years (mean (SD)) | 6.18 (8.90) | 10.54 (11.39) | 0.079 | 7.71 (7.68) | 0.551 | 4.85 (6.55) | 0.597 |
| missing/unknown | 0 (0) | 2 (7.69) |  | 2 (11.76) |  | 0 (0) |  |
| Year of index date (mean (SD)) | 2007.83 (3.79) | 2005.31 (4.19) | 0.010 | 2004.12 (3.30) | 0.001 | 2012.40 (2.92) | <0.001 |
| BMI kg/m<sup>2</sup> (mean (SD)) | 32.96 (2.84) | 32.24 (1.96) | 0.257 | 33.47 (3.15) | 0.534 | 32.52 (1.79) | 0.577 |
| High educational level (%) | 1 (2.1) | 1 (3.8) | 1.000 | 1 (5.9) | 1.000 | 1 (6.7) | 1.000 |
| missing/unknown | 22 (45.8) | 8 (30.8) |  | 4 (23.5) |  | 5 (33.3) |  |
| Ever smoker (%) | 15 (31.2) | 10 (38.5) | 0.712 | 3 (17.6) | 0.446 | 10 (66.7) | 0.032 |
| csDMARD at index date (%) | 39 (81.2) | 13 (50.0) | 0.011 | 15 (88.2) | 0.777 | 12 (80.0) | 1.000 |
| Prednisone at index date (%) | 24 (50.0) | 9 (34.6) | 0.305 | 12 (70.6) | 0.237 | 6 (40.0) | 0.703 |
| Seropositive (%) | 35 (72.9) | 18 (69.2) | 0.325 | 12 (70.6) | 1.000 | 14 (93.3) | 0.493 |
| missing/unknown | 5 (10.4) | 7 (26.9) |  | 3 (17.6) |  | 0 (0.0) |  |
| ESR (mean (SD)) | 20.82 (18.00) | 19.92 (15.29) | 0.833 | 25.94 (22.87) | 0.358 | 26.82 (24.51) | 0.362 |
| missing/unknown | 3 (6.25) | 1 (3.85) |  | 0 (0) |  | 4 (26.67) |  |
| CRP (mean (SD)) | 1.81 (1.41) | 1.88 (2.13) | 0.929 | 1.65 (1.91) | 0.882 | 1.27 (1.17) | 0.276 |
| missing/unknown | 30 (62.5) | 18 (69.23) |  | 15 (88.24) |  | 3 (20) |  |
| Tender joint counts 28 (mean (SD)) | 6.89 (7.69) | 7.65 (7.07) | 0.679 | 8.00 (6.75) | 0.602 | 10.23 (8.22) | 0.179 |
| missing/unknown | 2 (4.17) | 0 (0) |  | 0 (0) |  | 2 (13.33) |  |
| Swollen joint counts 28 (mean (SD)) | 6.54 (6.64) | 7.31 (6.30) | 0.634 | 8.41 (5.67) | 0.307 | 8.77 (5.46) | 0.273 |
| missing/unknown | 2 (4.17) | 0 (0) |  | 0 (0) |  | 2 (13.33) |  |
| Physician GDA (mean (SD)) | 4.42 (1.55) | 4.73 (2.41) | 0.649 | 5.55 (2.25) | 0.089 | 5.25 (2.14) | 0.185 |
| missing/unknown | 22 (45.83) | 15 (57.69) |  | 6 (35.29) |  | 3 (20) |  |
| DAS28-ESR (mean (SD)) | 4.05 (1.65) | 4.36 (1.52) | 0.435 | 4.56 (1.54) | 0.268 | 4.65 (1.78) | 0.287 |
| missing/unknown | 3 (6.25) | 1 (3.85) |  | 0 (0) |  | 4 (26.67) |  |
| DAS28-CRP (mean (SD)) | 4.45 (1.36) | 4.81 (1.02) | 0.509 | 3.55 (0.07) | 0.374 | 4.58 (1.26) | 0.789 |
| missing/unknown | 30 (62.5) | 18 (69.23) |  | 15 (88.24) |  | 3 (20) |  |
| RADAI-5 (mean (SD)) | 4.61 (2.15) | 4.80 (1.77) | 0.725 | 5.44 (2.09) | 0.195 | 4.12 (2.69) | 0.574 |
| missing/unknown | 6 (12.5) | 3 (11.5) |  | 1 (5.9) |  | 7 (46.7) |  |
| HAQ (mean (SD)) | 1.10 (0.65) | 1.17 (0.71) | 0.692 | 1.35 (0.80) | 0.221 | 0.61 (0.72) | 0.062 |
| missing/unknown | 6 (12.5) | 2 (7.69) |  | 0 (0) |  | 7 (46.67) |  |
| Osteoporosis | 4 (8.3) | 3 (11.5) | 0.973 | 1 (5.9) | 1.000 | 1 (6.7) | 1.000 |
| Fibromyalgia | 0 (0.0) | 0 (0.0) | NA | 0 (0.0) | NA | 0 (0.0) | NA |
| Other rheumatological disease | 17 (35.4) | 14 (53.8) | 0.198 | 9 (52.9) | 0.327 | 3 (20.0) | 0.423 |
| Psoriasis | 0 (0.0) | 1 (3.8) | 0.754 | 17 (100.0) | NA | 15 (100.0) | NA |
| Hyperlipidemia | 5 (10.4) | 2 (7.7) | 1.000 | 1 (5.9) | 0.946 | 6 (40.0) | 0.025 |
| Cardiac/cardiovascular event/disease | 21 (43.8) | 15 (57.7) | 0.367 | 8 (47.1) | 1.000 | 11 (73.3) | 0.088 |
| Cancer | 1 (2.1) | 1 (3.8) | 1.000 | 0 (0.0) | 1.000 | 2 (13.3) | 0.275 |
| Depression/anxiety | 4 (8.3) | 3 (11.5) | 0.973 | 3 (17.6) | 0.542 | 3 (20.0) | 0.433 |
| Diabetes | 6 (12.5) | 5 (19.2) | 0.664 | 1 (5.9) | 0.763 | 4 (26.7) | 0.365 |
| Fractures, surgeries,<br>musculoskeletal system | 3 (6.2) | 1 (3.8) | 1.000 | 2 (11.8) | 0.839 | 2 (13.3) | 0.735 |

Values are the number and column percentage, unless otherwise specified. Significance tests compare each drug of interest to Adalimumab, using chi-squared test for categorical variables, and t-test for continuous variables. For these tests, missing values did not function as a grouping variable.

Abbreviations: SD standard deviation; RA rheumatoid arthritis; BMI body mass index; csDMARD conventional synthetic disease modifying anti-rheumatic drug; ESR erythrocyte sedimentation rate; CRP C-reactive protein; DAS28-ESR 28-joint Disease Activity Score using erythrocyte sedimentation rate; DAS28-CRP 28-joint Disease Activity Score using C-reactive protein; RADAI-5 Rheumatoid Arthritis Disease Activity Index-Five; HAQ Health Assessment Questionnaire.

For additional information on seropositivity and comorbidities see footnote in Supplementary Table S3.

**Supplementary Table S8** Overweight female cohort, patient characteristics at baseline, stratified by first b/tsDMARD adalimumab, etanercept, infliximab, and abatacept.

| Overweight female | adalimumab<br>215 | etanercept<br>203 | p value | infliximab<br>91 | p value | abatacept<br>30 | p value |
| --- | --- | --- | --- | --- | --- | --- | --- |
| Age index (mean (SD)) | 56.84 (13.10) | 58.48 (12.58) | 0.192 | 57.29 (11.20) | 0.776 | 61.73 (11.82) | 0.054 |
| RA duration, years (mean (SD)) | 7.26 (8.00) | 8.94 (9.59) | 0.056 | 7.61 (7.61) | 0.726 | 6.36 (9.46) | 0.586 |
| missing/unknown | 10 (4.65) | 4 (1.97) |  | 1 (1.1) |  | 2 (6.67) |  |
| Year of index date (mean (SD)) | 2007.57 (3.44) | 2006.69 (4.93) | 0.034 | 2005.97 (4.66) | 0.001 | 2012.00 (2.82) | <0.001 |
| BMI kg/m2 (mean (SD)) | 27.16 (1.36) | 27.27 (1.35) | 0.434 | 27.13 (1.39) | 0.837 | 27.02 (1.37) | 0.602 |
| High educational level (%) | 10 (4.7) | 11 (5.4) | 0.686 | 6 (6.6) | 0.692 | 3 (10.0) | 0.487 |
| missing/unknown | 61 (28.4) | 73 (36.0) |  | 25 (27.5) |  | 7 (23.3) |  |
| Ever smoker (%) | 46 (21.4) | 43 (21.2) | 1.000 | 21 (23.1) | 0.862 | 12 (40.0) | 0.044 |
| csDMARD at index date (%) | 150 (69.8) | 129 (63.5) | 0.213 | 76 (83.5) | 0.018 | 23 (76.7) | 0.573 |
| Prednisone at index date (%) | 84 (39.1) | 85 (41.9) | 0.629 | 50 (54.9) | 0.015 | 7 (23.3) | 0.142 |
| Seropositive (%) | 158 (73.5) | 141 (69.5) | 0.703 | 72 (79.1) | 1.000 | 23 (76.7) | 1.000 |
| missing/unknown | 27 (12.6) | 31 (15.3) |  | 6 (6.6) |  | 2 (6.7) |  |
| ESR (mean (SD)) | 25.35 (18.20) | 25.59 (20.70) | 0.907 | 24.94 (19.45) | 0.866 | 26.55 (17.86) | 0.771 |
| missing/unknown | 28 (13.02) | 9 (4.43) |  | 7 (7.69) |  | 8 (26.67) |  |
| CRP (mean (SD)) | 1.75 (2.56) | 1.42 (1.91) | 0.357 | 1.15 (1.82) | 0.239 | 0.96 (1.12) | 0.149 |
| missing/unknown | 138 (64.19) | 123 (60.59) |  | 60 (65.93) |  | 6 (20) |  |
| Tender joint counts 28 (mean (SD)) | 7.44 (6.64) | 8.39 (8.08) | 0.206 | 7.34 (7.21) | 0.916 | 6.41 (6.18) | 0.447 |
| missing/unknown | 23 (10.7) | 5 (2.46) |  | 4 (4.4) |  | 3 (10) |  |
| Swollen joint counts 28 (mean (SD)) | 6.70 (5.16) | 6.87 (5.90) | 0.763 | 7.68 (5.68) | 0.158 | 5.56 (4.69) | 0.274 |
| missing/unknown | 22 (10.23) | 5 (2.46) |  | 4 (4.4) |  | 3 (10) |  |
| Physician GDA (mean (SD)) | 4.90 (2.22) | 5.06 (2.15) | 0.587 | 5.36 (1.85) | 0.204 | 3.87 (1.79) | 0.036 |
| missing/unknown | 84 (39.07) | 95 (46.8) |  | 44 (48.35) |  | 7 (23.33) |  |
| DAS28-ESR (mean (SD)) | 4.53 (1.19) | 4.47 (1.55) | 0.644 | 4.47 (1.42) | 0.725 | 4.52 (1.17) | 0.975 |
| missing/unknown | 29 (13.49) | 9 (4.43) |  | 7 (7.69) |  | 8 (26.67) |  |
| DAS28-CRP (mean (SD)) | 4.22 (1.17) | 4.33 (1.35) | 0.573 | 4.06 (1.17) | 0.521 | 4.04 (1.03) | 0.499 |
| missing/unknown | 138 (64.19) | 123 (60.59) |  | 60 (65.93) |  | 6 (20) |  |
| RADAI-5 (mean (SD)) | 4.97 (2.03) | 5.08 (2.14) | 0.647 | 5.30 (2.18) | 0.268 | 4.99 (2.13) | 0.976 |
| missing/unknown | 16 (7.4) | 10 (4.9) |  | 1 (1.1) |  | 2 (6.7) |  |
| HAQ (mean (SD)) | 1.17 (0.69) | 1.20 (0.75) | 0.757 | 1.27 (0.75) | 0.334 | 0.94 (0.69) | 0.149 |
| missing/unknown | 37 (17.21) | 21 (10.34) |  | 9 (9.89) |  | 10 (33.33) |  |
| Osteoporosis | 45 (20.9) | 51 (25.1) | 0.367 | 22 (24.2) | 0.634 | 7 (23.3) | 0.950 |
| Fibromyalgia | 0 (0.0) | 9 (4.4) | 0.005 | 5 (5.5) | 0.003 | 2 (6.7) | 0.007 |
| Other rheumatological disease | 70 (32.6) | 71 (35.0) | 0.675 | 29 (31.9) | 1.000 | 9 (30.0) | 0.942 |
| Psoriasis | 1 (0.5) | 2 (1.0) | 0.960 | 0 (0.0) | 1.000 | 1 (3.3) | 0.581 |
| Hyperlipidemia | 6 (2.8) | 14 (6.9) | 0.082 | 5 (5.5) | 0.409 | 2 (6.7) | 0.568 |
| Cardiac/cardiovascular event/disease | 77 (35.8) | 80 (39.4) | 0.511 | 35 (38.5) | 0.757 | 12 (40.0) | 0.807 |
| Cancer | 5 (2.3) | 9 (4.4) | 0.355 | 1 (1.1) | 0.798 | 1 (3.3) | 1.000 |
| Depression/anxiety | 26 (12.1) | 33 (16.3) | 0.280 | 20 (22.0) | 0.042 | 1 (3.3) | 0.261 |
| Diabetes | 12 (5.6) | 10 (4.9) | 0.936 | 6 (6.6) | 0.938 | 4 (13.3) | 0.224 |
| Fractures, surgeries<br>musculoskeletal system | 13 (6.0) | 28 (13.8) | 0.013 | 6 (6.6) | 1.000 | 1 (3.3) | 0.857 |

Values are the number and column percentage, unless otherwise specified. Significance tests compare each drug of interest to Adalimumab, using chi-squared test for categorical variables, and t-test for continuous variables. For these tests, missing values did not function as a grouping variable.

Abbreviations: SD standard deviation; RA rheumatoid arthritis; BMI body mass index; csDMARD conventional synthetic disease modifying anti-rheumatic drug; ESR erythrocyte sedimentation rate; CRP C-reactive protein; DAS28-ESR 28-joint Disease Activity Score using erythrocyte sedimentation rate; DAS28-CRP 28-joint Disease Activity Score using C-reactive protein; RADAI-5 Rheumatoid Arthritis Disease Activity Index-Five; HAQ Health Assessment Questionnaire.

For additional information on seropositivity and comorbidities see footnote in Supplementary Table S3.

**Supplementary Table S9** Overweight male cohort, patient characteristics at baseline, stratified by first b/tsDMARD adalimumab, etanercept, infliximab, and abatacept.

| Overweight male | adalimumab<br>121 | etanercept<br>93 | p value | infliximab<br>59 | p value | abatacept<br>17 | p value |
| --- | --- | --- | --- | --- | --- | --- | --- |
| Age index (mean (SD)) | 58.05 (11.43) | 56.35 (11.64) | 0.285 | 56.22 (11.64) | 0.318 | 65.22 (8.63) | 0.014 |
| RA duration, years (mean (SD)) | 6.83 (7.35) | 7.35 (8.26) | 0.628 | 7.74 (8.86) | 0.475 | 7.01 (6.58) | 0.924 |
| missing/unknown | 2 (1.65) | 3 (3.23) |  | 3 (5.08) |  | 1 (5.88) |  |
| Year of index date (mean (SD)) | 2007.88 (3.77) | 2006.77 (5.24) | 0.073 | 2005.86 (4.78) | 0.002 | 2012.88 (2.12) | <0.001 |
| BMI kg/m2 (mean (SD)) | 27.16 (1.33) | 27.09 (1.27) | 0.698 | 27.14 (1.36) | 0.931 | 27.49 (1.47) | 0.349 |
| High educational level (%) | 19 (15.7) | 10 (10.8) | 0.505 | 6 (10.2) | 0.354 | 3 (17.6) | 1.000 |
| missing/unknown | 37 (30.6) | 33 (35.5) |  | 16 (27.1) |  | 4 (23.5) |  |
| Ever smoker (%) | 48 (39.7) | 33 (35.5) | 0.629 | 22 (37.3) | 0.885 | 10 (58.8) | 0.217 |
| csDMARD at index date (%) | 76 (62.8) | 60 (64.5) | 0.909 | 50 (84.7) | 0.004 | 11 (64.7) | 1.000 |
| Prednisone at index date (%) | 50 (41.3) | 48 (51.6) | 0.174 | 34 (57.6) | 0.058 | 6 (35.3) | 0.833 |
| Seropositive (%) | 86 (71.1) | 70 (75.3) | 1.000 | 47 (79.7) | 0.719 | 14 (82.4) | 0.261 |
| missing/unknown | 20 (16.5) | 10 (10.8) |  | 6 (10.2) |  | 3 (17.6) |  |
| ESR (mean (SD)) | 23.93 (21.76) | 26.53 (26.71) | 0.459 | 26.18 (24.18) | 0.543 | 32.19 (18.41) | 0.151 |
| missing/unknown | 11 (9.09) | 12 (12.9) |  | 2 (3.39) |  | 1 (5.88) |  |
| CRP (mean (SD)) | 2.68 (4.05) | 2.38 (4.80) | 0.752 | 1.24 (1.45) | 0.146 | 1.78 (1.81) | 0.378 |
| missing/unknown | 70 (57.85) | 58 (62.37) |  | 41 (69.49) |  | 0 (0) |  |
| Tender joint counts 28 (mean (SD)) | 7.75 (7.35) | 6.94 (6.64) | 0.427 | 6.83 (6.32) | 0.419 | 5.31 (5.47) | 0.205 |
| missing/unknown | 10 (8.26) | 7 (7.53) |  | 1 (1.69) |  | 1 (5.88) |  |
| Swollen joint counts 28 (mean (SD)) | 7.25 (6.08) | 6.24 (6.01) | 0.247 | 8.28 (6.45) | 0.312 | 6.24 (6.00) | 0.520 |
| missing/unknown | 11 (9.09) | 7 (7.53) |  | 1 (1.69) |  | 0 (0) |  |
| Physician GDA (mean (SD)) | 4.97 (2.38) | 4.54 (2.08) | 0.315 | 5.87 (2.32) | 0.086 | 4.82 (1.94) | 0.814 |
| missing/unknown | 52 (42.98) | 45 (48.39) |  | 29 (49.15) |  | 0 (0) |  |
| DAS28-ESR (mean (SD)) | 4.36 (1.56) | 4.31 (1.39) | 0.829 | 4.35 (1.58) | 0.962 | 4.44 (1.14) | 0.848 |
| missing/unknown | 12 (9.92) | 12 (12.9) |  | 2 (3.39) |  | 1 (5.88) |  |
| DAS28-CRP (mean (SD)) | 4.37 (1.26) | 3.86 (1.14) | 0.061 | 3.92 (1.46) | 0.217 | 3.89 (1.16) | 0.182 |
| missing/unknown | 71 (58.68) | 58 (62.37) |  | 41 (69.49) |  | 1 (5.88) |  |
| RADAI-5 (mean (SD)) | 4.58 (2.16) | 4.85 (2.26) | 0.428 | 4.55 (2.12) | 0.937 | 4.68 (2.76) | 0.881 |
| missing/unknown | 30 (24.8) | 18 (19.4) |  | 6 (10.2) |  | 5 (29.4) |  |
| HAQ (mean (SD)) | 0.90 (0.70) | 0.97 (0.68) | 0.514 | 1.20 (0.69) | 0.011 | 0.93 (0.72) | 0.875 |
| missing/unknown | 26 (21.49) | 17 (18.28) |  | 5 (8.47) |  | 4 (23.53) |  |
| Osteoporosis | 14 (11.6) | 10 (10.8) | 1.000 | 11 (18.6) | 0.290 | 4 (23.5) | 0.324 |
| Fibromyalgia | 0 (0) | 0 (0) | NA | 0 (0) | NA | 0 (0) | NA |
| Other rheumatological disease | 31 (25.6) | 26 (28.0) | 0.820 | 16 (27.1) | 0.973 | 4 (23.5) | 1.000 |
| Psoriasis | 1 (0.8) | 1 (1.1) | 1.000 | 0 (0.0) | 1.000 | 0 (0.0) | 1.000 |
| Hyperlipidemia | 10 (8.3) | 11 (11.8) | 0.524 | 3 (5.1) | 0.641 | 5 (29.4) | 0.027 |
| Cardiac/cardiovascular event/disease | 46 (38.0) | 37 (39.8) | 0.903 | 23 (39.0) | 1.000 | 9 (52.9) | 0.362 |
| Cancer | 1 (0.8) | 4 (4.3) | 0.226 | 0 (0.0) | 1.000 | 1 (5.9) | 0.583 |
| Depression/anxiety | 5 (4.1) | 9 (9.7) | 0.178 | 7 (11.9) | 0.102 | 1 (5.9) | 1.000 |
| Diabetes | 14 (11.6) | 9 (9.7) | 0.825 | 2 (3.4) | 0.126 | 4 (23.5) | 0.324 |
| Fractures, surgeries,<br>musculoskeletal system | 13 (10.7) | 6 (6.5) | 0.394 | 7 (11.9) | 1.000 | 1 (5.9) | 0.847 |

Values are the number and column percentage, unless otherwise specified. Significance tests compare each drug of interest to Adalimumab, using chi-squared test for categorical variables, and t-test for continuous variables. For these tests, missing values did not function as a grouping variable.

Abbreviations: SD standard deviation; RA rheumatoid arthritis; BMI body mass index; csDMARD conventional synthetic disease modifying anti-rheumatic drug; ESR erythrocyte sedimentation rate; CRP C-reactive protein; DAS28-ESR 28-joint Disease Activity Score using erythrocyte sedimentation rate; DAS28-CRP 28-joint Disease Activity Score using C-reactive protein; RADAI-5 Rheumatoid Arthritis Disease Activity Index-Five; HAQ Health Assessment Questionnaire.

For additional information on seropositivity and comorbidities see footnote in Supplementary Table S3.

**Supplementary Table S10** Normal weight female cohort, patient characteristics at baseline, stratified by first b/tsDMARD adalimumab, etanercept, infliximab, and abatacept.

| Normal weight female | adalimumab<br>365 | etanercept<br>393 | p value | infliximab<br>207 | p value | abatacept<br>51 | p value |
| --- | --- | --- | --- | --- | --- | --- | --- |
| Age index (mean (SD)) | 53.03 (13.95) | 53.84 (14.78) | 0.438 | 53.70 (14.18) | 0.583 | 61.81 (13.39) | <0.001 |
| RA duration, years (mean (SD)) | 8.86 (9.45) | 8.96 (9.31) | 0.894 | 10.49 (9.72) | 0.054 | 9.99 (10.98) | 0.439 |
| missing/unknown | 9 (2.47) | 9 (2.29) |  | 5 (2.42) |  | 1 (1.96) |  |
| Year of index date (mean (SD)) | 2007.10 (3.20) | 2005.84 (4.84) | <0.001 | 2004.61 (4.11) | <0.001 | 2012.78 (2.80) | <0.001 |
| BMI kg/m2 (mean (SD)) | 22.08 (1.76) | 21.97 (1.78) | 0.365 | 22.07 (1.81) | 0.963 | 22.24 (1.70) | 0.556 |
| High educational level (%) | 38 (10.4) | 28 (7.1) | 0.191 | 17 (8.2) | 0.708 | 6 (11.8) | 1.000 |
| missing/unknown | 113 (31.0) | 134 (34.1) |  | 77 (37.2) |  | 10 (19.6) |  |
| Ever smoker (%) | 91 (24.9) | 80 (20.4) | 0.156 | 37 (17.9) | 0.066 | 14 (27.5) | 0.829 |
| csDMARD at index date (%) | 260 (71.2) | 222 (56.5) | <0.001 | 167 (80.7) | 0.017 | 33 (64.7) | 0.428 |
| Prednisone at index date (%) | 145 (39.7) | 160 (40.7) | 0.839 | 100 (48.3) | 0.057 | 22 (43.1) | 0.754 |
| Seropositive (%) | 266 (72.9) | 308 (78.4) | 0.401 | 169 (81.6) | 0.013 | 39 (76.5) | 0.376 |
| missing/unknown | 49 (13.4) | 38 (9.7) |  | 24 (11.6) |  | 1 (2.0) |  |
| ESR (mean (SD)) | 23.20 (19.08) | 24.74 (22.92) | 0.347 | 28.03 (24.61) | 0.014 | 25.87 (18.95) | 0.416 |
| missing/unknown | 47 (12.88) | 39 (9.92) |  | 21 (10.14) |  | 13 (25.49) |  |
| CRP (mean (SD)) | 1.24 (1.49) | 1.21 (1.62) | 0.899 | 1.45 (2.05) | 0.487 | 1.49 (2.05) | 0.417 |
| missing/unknown | 256 (70.14) | 285 (72.52) |  | 162 (78.26) |  | 12 (23.53) |  |
| Tender joint counts 28 (mean (SD)) | 6.77 (6.42) | 6.86 (6.79) | 0.864 | 6.66 (6.51) | 0.847 | 6.98 (5.80) | 0.842 |
| missing/unknown | 33 (9.04) | 22 (5.6) |  | 15 (7.25) |  | 7 (13.73) |  |
| Swollen joint counts 28 (mean (SD)) | 6.66 (5.70) | 6.75 (6.05) | 0.849 | 8.74 (6.92) | <0.001 | 6.62 (5.54) | 0.965 |
| missing/unknown | 34 (9.32) | 22 (5.6) |  | 14 (6.76) |  | 6 (11.76) |  |
| Physician GDA (mean (SD)) | 5.04 (2.11) | 5.14 (2.21) | 0.624 | 5.15 (2.04) | 0.646 | 4.46 (1.79) | 0.128 |
| missing/unknown | 175 (47.95) | 192 (48.85) |  | 103 (49.76) |  | 16 (31.37) |  |
| DAS28-ESR (mean (SD)) | 4.33 (1.34) | 4.28 (1.45) | 0.644 | 4.51 (1.48) | 0.160 | 4.48 (0.97) | 0.507 |
| missing/unknown | 48 (13.15) | 39 (9.92) |  | 22 (10.63) |  | 14 (27.45) |  |
| DAS28-CRP (mean (SD)) | 4.11 (1.17) | 4.05 (1.09) | 0.683 | 4.03 (1.23) | 0.716 | 4.13 (1.15) | 0.929 |
| missing/unknown | 257 (70.41) | 285 (72.52) |  | 163 (78.74) |  | 13 (25.49) |  |
| RADAI-5 (mean (SD)) | 4.62 (2.17) | 4.67 (2.16) | 0.788 | 4.65 (2.05) | 0.888 | 4.21 (1.66) | 0.264 |
| missing/unknown | 73 (20.00) | 55 (13.99) |  | 41 (19.81) |  | 14 (27.45) |  |
| HAQ (mean (SD)) | 0.99 (0.72) | 1.05 (0.73) | 0.263 | 1.21 (0.70) | 0.001 | 0.76 (0.57) | 0.056 |
| missing/unknown | 65 (17.81) | 51 (12.98) |  | 26 (12.56) |  | 12 (23.53) |  |
| Osteoporosis | 78 (21.4) | 87 (22.1) | 0.867 | 51 (24.6) | 0.427 | 22 (43.1) | 0.001 |
| Fibromyalgia | 0 (0.0) | 1 (0.3) | 1.000 | 3 (1.4) | 0.088 | 0 (0) | NA |
| Other rheumatological disease | 82 (22.5) | 99 (25.2) | 0.427 | 59 (28.5) | 0.131 | 14 (27.5) | 0.539 |
| Psoriasis | 4 (1.1) | 1 (0.3) | 0.327 | 1 (0.5) | 0.772 | 0 (0.0) | 1.000 |
| Hyperlipidemia | 6 (1.6) | 10 (2.5) | 0.542 | 1 (0.5) | 0.414 | 6 (11.8) | <0.001 |
| Cardiac/cardiovascular event/disease | 60 (16.4) | 89 (22.6) | 0.040 | 44 (21.3) | 0.186 | 26 (51.0) | <0.001 |
| Cancer | 3 (0.8) | 8 (2.0) | 0.275 | 4 (1.9) | 0.444 | 3 (5.9) | 0.027 |
| Depression/anxiety | 27 (7.4) | 40 (10.2) | 0.223 | 25 (12.1) | 0.085 | 6 (11.8) | 0.421 |
| Diabetes | 2 (0.5) | 9 (2.3) | 0.089 | 2 (1.0) | 0.956 | 3 (5.9) | 0.010 |
| Fractures, surgeries, musculoskeletal system | 40 (11.0) | 51 (13.0) | 0.458 | 33 (15.9) | 0.113 | 2 (3.9) | 0.189 |

Values are the number and column percentage, unless otherwise specified. Significance tests compare each drug of interest to Adalimumab, using chi-squared test for categorical variables, and t-test for continuous variables. For these tests, missing values did not function as a grouping variable.

Abbreviations: SD standard deviation; RA rheumatoid arthritis; BMI body mass index; csDMARD conventional synthetic disease modifying anti-rheumatic drug; ESR erythrocyte sedimentation rate; CRP C-reactive protein; DAS28-ESR 28-joint Disease Activity Score using erythrocyte sedimentation rate; DAS28-CRP 28-joint Disease Activity Score using C-reactive protein; RADAI-5 Rheumatoid Arthritis Disease Activity Index-Five; HAQ Health Assessment Questionnaire.

For additional information on seropositivity and comorbidities see footnote in Supplementary Table S3.

**Supplementary Table S11** Normal weight male cohort, patient characteristics at baseline, stratified by first b/tsDMARD adalimumab, etanercept, infliximab, and abatacept.

| Normal weight male | adalimumab<br>77 | etanercept<br>89 | p value | infliximab<br>52 | p value | abatacept<br>9 | p value |
| --- | --- | --- | --- | --- | --- | --- | --- |
| Age index (mean (SD)) | 54.24 (12.98) | 56.40 (14.14) | 0.309 | 50.97 (14.60) | 0.185 | 62.91 (19.86) | 0.078 |
| RA duration, years (mean (SD)) | 5.57 (7.12) | 7.31 (7.06) | 0.122 | 7.15 (6.95) | 0.220 | 3.91 (4.36) | 0.521 |
| missing/unknown | 1 (1.3) | 4 (4.49) |  | 1 (1.92) |  | 1 (11.11) |  |
| Year of index date (mean (SD)) | 2008.47 (4.32) | 2005.73 (5.16) | <0.001 | 2005.58 (3.70) | <0.001 | 2012.33 (1.94) | 0.010 |
| BMI kg/m2 (mean (SD)) | 22.58 (1.76) | 22.55 (1.90) | 0.897 | 22.57 (1.81) | 0.965 | 23.55 (1.61) | 0.120 |
| High educational level (%) | 12 (15.6) | 19 (21.3) | 0.430 | 7 (13.5) | 0.977 | 0 (0.0) | 0.278 |
| missing/unknown | 27 (35.1) | 31 (34.8) |  | 19 (36.5) |  | 1 (11.1) |  |
| Ever smoker (%) | 31 (40.3) | 39 (43.8) | 0.760 | 18 (34.6) | 0.643 | 4 (44.4) | 1.000 |
| csDMARD at index date (%) | 56 (72.7) | 43 (48.3) | 0.002 | 41 (78.8) | 0.561 | 5 (55.6) | 0.493 |
| Prednisone at index date (%) | 23 (29.9) | 33 (37.1) | 0.415 | 24 (46.2) | 0.089 | 5 (55.6) | 0.238 |
| Seropositive (%) | 59 (76.6) | 71 (79.8) | 0.214 | 39 (75.0) | 0.708 | 7 (77.8) | 1.000 |
| missing/unknown | 7 (9.1) | 12 (13.5) |  | 8 (15.4) |  | 1 (11.1) |  |
| ESR (mean (SD)) | 24.89 (24.56) | 30.08 (23.86) | 0.193 | 18.84 (23.48) | 0.185 | 31.62 (35.13) | 0.488 |
| missing/unknown | 11 (14.29) | 4 (4.49) |  | 3 (5.77) |  | 1 (11.11) |  |
| CRP (mean (SD)) | 1.42 (1.84) | 1.78 (1.54) | 0.426 | 1.40 (1.84) | 0.979 | 1.36 (1.18) | 0.933 |
| missing/unknown | 43 (55.84) | 65 (73.03) |  | 38 (73.08) |  | 1 (11.11) |  |
| Tender joint counts 28 (mean (SD)) | 6.34 (6.39) | 5.96 (6.03) | 0.706 | 6.31 (7.66) | 0.977 | 3.50 (3.02) | 0.220 |
| missing/unknown | 7 (9.09) | 4 (4.49) |  | 3 (5.77) |  | 1 (11.11) |  |
| Swollen joint counts 28 (mean (SD)) | 6.10 (5.56) | 7.19 (6.26) | 0.254 | 7.20 (6.85) | 0.330 | 3.62 (2.88) | 0.221 |
| missing/unknown | 5 (6.49) | 4 (4.49) |  | 2 (3.85) |  | 1 (11.11) |  |
| Physician GDA (mean (SD)) | 4.75 (2.18) | 5.12 (2.03) | 0.377 | 5.58 (2.70) | 0.155 | 4.14 (1.68) | 0.482 |
| missing/unknown | 25 (32.47) | 38 (42.7) |  | 28 (53.85) |  | 2 (22.22) |  |
| DAS28-ESR (mean (SD)) | 4.07 (1.64) | 4.36 (1.34) | 0.241 | 3.71 (1.73) | 0.263 | 3.64 (1.82) | 0.488 |
| missing/unknown | 12 (15.58) | 5 (5.62) |  | 4 (7.69) |  | 1 (11.11) |  |
| DAS28-CRP (mean (SD)) | 4.12 (1.38) | 4.59 (1.25) | 0.193 | 3.79 (1.09) | 0.455 | 3.49 (1.26) | 0.249 |
| missing/unknown | 44 (57.14) | 66 (74.16) |  | 39 (75) |  | 1 (11.11) |  |
| RADAI-5 (mean (SD)) | 4.56 (2.46) | 4.79 (2.04) | 0.550 | 4.36 (2.28) | 0.654 | 5.03 (2.49) | 0.638 |
| missing/unknown | 15 (19.5) | 8 (9) |  | 4 (7.7) |  | 2 (22.2) |  |
| HAQ (mean (SD)) | 0.85 (0.63) | 0.96 (0.64) | 0.326 | 0.77 (0.67) | 0.496 | 0.89 (0.97) | 0.882 |
| missing/unknown | 15 (19.48) | 8 (8.99) |  | 3 (5.77) |  | 2 (22.22) |  |
| Osteoporosis | 10 (13.0) | 14 (15.7) | 0.780 | 10 (19.2) | 0.476 | 1 (11.1) | 1.000 |
| Fibromyalgia | 1 (1.3) | 0 (0.0) | 0.942 | 1 (1.9) | 1.000 | 0 (0.0) | 1.000 |
| Other rheumatological disease | 17 (22.1) | 19 (21.3) | 1.000 | 11 (21.2) | 1.000 | 2 (22.2) | 1.000 |
| Psoriasis | 1 (1.3) | 1 (1.1) | 1.000 | 1 (1.9) | 1.000 | 0 (0.0) | 1.000 |
| Hyperlipidemia | 3 (3.9) | 4 (4.5) | 1.000 | 2 (3.8) | 1.000 | 1 (11.1) | 0.892 |
| Cardiac/cardiovascular event/disease | 18 (23.4) | 26 (29.2) | 0.501 | 12 (23.1) | 1.000 | 5 (55.6) | 0.096 |
| Cancer | 1 (1.3) | 1 (1.1) | 1.000 | 2 (3.8) | 0.729 | 1 (11.1) | 0.497 |
| Depression/anxiety | 3 (3.9) | 6 (6.7) | 0.643 | 5 (9.6) | 0.343 | 0 (0.0) | 1.000 |
| Diabetes | 4 (5.2) | 12 (13.5) | 0.123 | 2 (3.8) | 1.000 | 1 (11.1) | 1.000 |
| Fractures, surgeries,<br>musculoskeletal system | 3 (3.9) | 9 (10.1) | 0.214 | 5 (9.6) | 0.343 | 1 (11.1) | 0.892 |

Values are the number and column percentage, unless otherwise specified. Significance tests compare each drug of interest to Adalimumab, using chi-squared test for categorical variables, and t-test for continuous variables. For these tests, missing values did not function as a grouping variable.

Abbreviations: SD standard deviation; RA rheumatoid arthritis; BMI body mass index; csDMARD conventional synthetic disease modifying anti-rheumatic drug; ESR erythrocyte sedimentation rate; CRP C-reactive protein; DAS28-ESR 28-joint Disease Activity Score using erythrocyte sedimentation rate; DAS28-CRP 28-joint Disease Activity Score using C-reactive protein; RADAI-5 Rheumatoid Arthritis Disease Activity Index-Five; HAQ Health Assessment Questionnaire.

For additional information on seropositivity and comorbidities see footnote in Supplementary Table S3.

**Supplementary Table S12** Sensitivity analyses excluding patients who miss information on outcome during follow-up (EPMOI, Excluding Patients Missing Outcome Information), overall BMI cohorts.

| Sensitivity analyses (EPMOI) |  |  |  |  |
| --- | --- | --- | --- | --- |
|  |  | n all | n event | OR |
| Obese - overall | **DAS28-remission** |  |  |  |
|  | adalimumab | 71 | 25 | 1 (ref.) |
|  | etanercept | 64 | 21 | 1.01 (0.49-2.11) |
|  | infliximab | 32 | 7 | 0.49 (0.17-1.26) |
|  | abatacept | 18 | 6 | 0.91 (0.28-2.73) |
|  | **DAS28-LDA** |  |  |  |
|  | adalimumab | 71 | 37 | 1 (ref.) |
|  | etanercept | 64 | 32 | 0.95 (0.48-1.89) |
|  | infliximab | 32 | 15 | 0.85 (0.36-1.98) |
|  | abatacept | 18 | 8 | 0.68 (0.23-1.95) |
|  | **RADAI-5-remission** |  |  |  |
|  | adalimumab | 63 | 11 | 1 (ref.) |
| Overweight - overall | etanercept | 56 | 9 | 0.95 (0.35-2.53) |
|  | infliximab | 28 | 5 | 1.01 (0.29-3.14) |
|  | abatacept | 20 | 4 | 1.21 (0.30-4.13) |
|  | **DAS28-remission** |  |  |  |
|  | adalimumab | 147 | 55 | 1 (ref.) |
|  | etanercept | 136 | 44 | 0.83 (0.50-1.36) |
|  | infliximab | 81 | 33 | 1.15 (0.65-2.02) |
|  | abatacept | 23 | 8 | 1.18 (0.44-2.98) |
|  | **DAS28-LDA** |  |  |  |
|  | adalimumab | 147 | 84 | 1 (ref.) |
|  | etanercept | 136 | 62 | 0.63 (0.39-1.02) |
|  | infliximab | 81 | 44 | 0.89 (0.51-1.54) |
|  | abatacept | 23 | 13 | 1.15 (0.47-2.91) |
| Normal weight - overall | **RADAI-5-remission** |  |  |  |
|  | adalimumab | 133 | 35 | 1 (ref.) |
|  | etanercept | 121 | 16 | 0.43 (0.22-0.82) |
|  | infliximab | 80 | 18 | 0.82 (0.42-1.55) |
|  | abatacept | 13 | 4 | 1.33 (0.34-4.47) |
|  | **DAS28-remission** |  |  |  |
|  | adalimumab | 204 | 85 | 1 (ref.) |
|  | etanercept | 256 | 99 | 0.89 (0.61-1.31) |
|  | infliximab | 147 | 55 | 0.82 (0.52-1.28) |
|  | abatacept | 29 | 14 | 1.82 (0.81-4.10) |
|  | **DAS28-LDA** |  |  |  |
|  | adalimumab | 204 | 122 | 1 (ref.) |
|  | etanercept | 256 | 148 | 0.93 (0.63-1.37) |
|  | infliximab | 147 | 82 | 0.84 (0.54-1.30) |
|  | abatacept | 29 | 20 | 2.05 (0.89-5.04) |
|  | **RADAI-5-remission** |  |  |  |
|  | adalimumab | 186 | 49 | 1 (ref.) |
|  | etanercept | 243 | 69 | 1.15 (0.75-1.77) |
|  | infliximab | 134 | 38 | 1.11 (0.67-1.83) |
|  | abatacept | 30 | 9 | 1.36 (0.55-3.13) |

OR odds ratio adjusted for sex and age.

Abbreviations: EPMOI Excluding Patients Missing Outcome Information; n number; ref reference; DAS28-remisison 28-joint Disease Activity Score remission; RADAI-5 Rheumatoid Arthritis Disease Activity Index-Five.

**Supplementary Table S13** Sensitivity analyses excluding patients who miss information on outcome during follow-up (EPMOI, Excluding Patients Missing Outcome Information), female and male BMI cohorts.

|  |  | Sensitivity analyses (EPMOI) |  |  |  |  | Sensitivity analyses (EPMOI) |  |  |
| --- | --- | --- | --- | --- | --- | --- | --- | --- | --- |
|  |  | n<br>all | n<br>event | OR |  |  | n<br>all | n<br>event | OR |
| Obese - female | **DAS28-remission** |  |  |  | Obese - male |  |  |  |  |
|  | adalimumab | 50 | 16 | 1 (ref.) |  | adalimumab | 21 | 9 | 1 (ref.) |
|  | etanercept | 54 | 16 | 0.88 (0.38-2.03) |  | etanercept | 10 | 5 | 1.36 (0.29-6.51) |
|  | infliximab | 22 | 2 | 0.20 (0.03-0.79) |  | infliximab | 10 | 5 | 1.35 (0.29-6.38) |
|  | abatacept | 12 | 4 | 1.15 (0.27-4.32) |  | abatacept | 6 | 2 | 0.67 (0.08-4.25) |
|  | **DAS28-LDA** |  |  |  |  | **DAS28-LDA** |  |  |  |
|  | adalimumab | 50 | 27 | 1 (ref.) |  | adalimumab | 21 | 10 | 1 (ref.) |
|  | etanercept | 54 | 26 | 0.81 (0.37-1.76) |  | etanercept | 10 | 6 | 1.54 (0.33-7.76) |
|  | infliximab | 22 | 8 | 0.53 (0.18-1.48) |  | infliximab | 10 | 7 | 2.47 (0.52-14.08) |
|  | abatacept | 12 | 6 | 0.78 (0.21-2.84) |  | abatacept | 6 | 2 | 0.55 (0.07-3.51) |
|  | **RADAI-5-remission** |  |  |  |  | **RADAI-5-remission** |  |  |  |
|  | adalimumab | 44 | 7 | 1 (ref.) |  | adalimumab | 19 | 4 | 1 (ref.) |
| Overweight - female | etanercept | 47 | 8 | 1.08 (0.35-3.38) | Overweight - male | etanercept | 9 | 1 | 0.48 (0.02-4.04) |
|  | infliximab | 19 | 2 | 0.60 (0.08-2.84) |  | infliximab | 9 | 3 | 1.91 (0.3-11.65) |
|  | abatacept | 14 | 3 | 1.49 (0.28-6.51) |  | abatacept | 6 | 1 | 0.75 (0.03-6.83) |
|  | **DAS28-remission** |  |  |  |  | **DAS28-remission** |  |  |  |
|  | adalimumab | 92 | 31 | 1 (ref.) |  | adalimumab | 55 | 24 | 1 (ref.) |
|  | etanercept | 96 | 31 | 1.02 (0.55-1.90) |  | etanercept | 40 | 13 | 0.6 (0.25-1.40) |
|  | infliximab | 52 | 23 | 1.65 (0.81-3.37) |  | infliximab | 29 | 10 | 0.64 (0.24-1.63) |
|  | abatacept | 19 | 8 | 1.96 (0.68-5.58) |  | abatacept | 4 | 0 | - |
|  | **DAS28-LDA** |  |  |  |  | **DAS28-LDA** |  |  |  |
|  | adalimumab | 92 | 53 | 1 (ref.) |  | adalimumab | 55 | 31 | 1 (ref.) |
|  | etanercept | 96 | 45 | 0.68 (0.38-1.23) |  | etanercept | 40 | 17 | 0.56 (0.24-1.28) |
|  | infliximab | 52 | 28 | 0.88 (0.44-1.76) |  | infliximab | 29 | 16 | 0.93 (0.37-2.33) |
| Normal weight - female | abatacept | 19 | 12 | 1.58 (0.57-4.69) |  | abatacept | 4 | 1 | 0.29 (0.01-2.53) |
|  | **RADAI-5-remission** |  |  |  |  | **RADAI-5-remission** |  |  |  |
|  | adalimumab | 85 | 19 | 1 (ref.) |  | adalimumab | 48 | 16 | 1 (ref.) |
|  | etanercept | 86 | 11 | 0.51 (0.22-1.15) |  | etanercept | 35 | 5 | 0.33 (0.10-0.96) |
|  | infliximab | 52 | 13 | 1.16 (0.51-2.60) |  | infliximab | 28 | 5 | 0.44 (0.13-1.29) |
|  | abatacept | 11 | 4 | 2.04 (0.49-7.61) |  | abatacept | 2 | 0 | - |
|  | **DAS28-remission** |  |  |  | Normal weight - male | **DAS28-remission** |  |  |  |
|  | adalimumab | 170 | 69 | 1 (ref.) |  | adalimumab | 34 | 16 | 1 (ref.) |
|  | etanercept | 212 | 78 | 0.85 (0.56-1.30) |  | etanercept | 44 | 21 | 1.25 (0.48-3.33) |
|  | infliximab | 124 | 42 | 0.76 (0.46-1.23) |  | infliximab | 23 | 13 | 1.03 (0.33-3.27) |
|  | abatacept | 25 | 13 | 1.99 (0.84-4.77) |  | abatacept | 4 | 1 | 1.17 (0.05-12.24) |
|  | **DAS28-LDA** |  |  |  |  | **DAS28-LDA** |  |  |  |
|  | adalimumab | 170 | 96 | 1 (ref.) |  | adalimumab | 34 | 26 | 1 (ref.) |
|  | etanercept | 212 | 119 | 0.99 (0.66-1.50) |  | etanercept | 44 | 29 | 0.67 (0.23-1.87) |
|  | infliximab | 124 | 66 | 0.89 (0.55-1.43) |  | infliximab | 23 | 16 | 0.51 (0.14-1.81) |
|  | abatacept | 25 | 17 | 2.09 (0.86-5.46) |  | abatacept | 4 | 3 | 2.15 (0.21-50.93) |
|  | **RADAI-5-remission** |  |  |  |  | **RADAI-5-remission** |  |  |  |
|  | adalimumab | 157 | 43 | 1 (ref.) |  | adalimumab | 29 | 6 | 1 (ref.) |
|  | etanercept | 203 | 60 | 1.14 (0.72-1.82) |  | etanercept | 40 | 9 | 1.21 (0.37-4.14) |
|  | infliximab | 114 | 32 | 1.05 (0.61-1.81) |  | infliximab | 20 | 6 | 1.55 (0.40-5.96) |
|  | abatacept | 27 | 7 | 1.04 (0.38-2.57) |  | abatacept | 3 | 2 | 10.81 (0.77-289.43) |

OR odds ratio adjusted for age.

Abbreviations: EPMOI Excluding Patients Missing Outcome Information; n number; ref reference; DAS28-remisison 28-joint Disease Activity Score remission; RADAI-5 Rheumatoid Arthritis Disease Activity Index-Five.

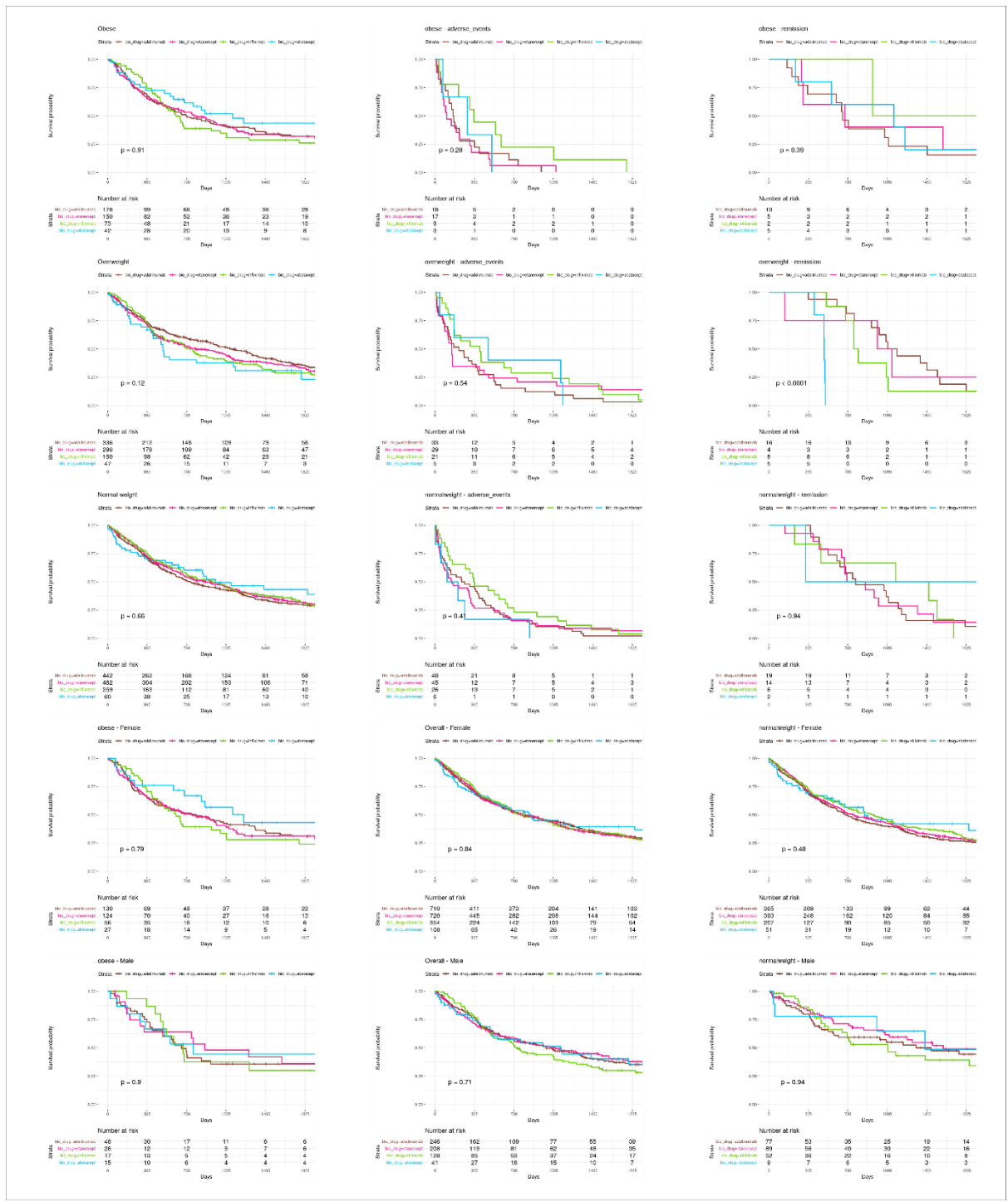

**Supplementary Figure S4** Kaplan-Meier curves for drug survival in the BMI cohorts overall and stratifying by sex, and further stratifying by reason of treatment stop as adverse event(s) or remission as recorded by the clinician. Log-rank p value indicated in each graph, and frequency table described below each respective graph.
